# Deep learning enables diagnosis of atrial cardiomyopathy from routine 12-lead electrocardiogram

**DOI:** 10.64898/2026.01.12.26343962

**Authors:** Julian Deseoe, Ezequiel de la Rosa, Martin Haensel, Lisa Herzog, Andreas R. Luft, Beate Sick, Jan Steffel, Alexander Breitenstein, Gregory Y. H. Lip, Björn Menze, Susanne Wegener

## Abstract

**Background and Aims:** Atrial cardiomyopathy (AtCM) contributes to the development of atrial fibrillation (AF), heart failure (HF) and stroke. Imaging-derived measures of left atrial (LA) structure and function are used to diagnose AtCM. Considering the tight coupling of heart structure and rhythm generation, this information might also be derived from 12-lead electrocardiogram (ECG), which is low-cost and readily available.

**Methods:** We finetuned a deep learning (DL) ECG foundational model to predict LA imaging indices based on 26’134 ECGs from the UK Biobank cohort. We then investigated if the ECG-predicted imaging features improved risk stratification of AF beyond the CHARGE-AF Score on a test set from the UK Biobank. We repeated this analysis for diagnosis of HF. We then externally validated our model and applied it to a cohort of stroke patients.

**Results:** Our DL model successfully predicted LA imaging indices from 12 lead ECG. In the UK Biobank test set, the ECG-predicted LA imaging features significantly improved risk stratification for AF beyond the CHARGE-AF score. ECG-predicted imaging markers showed superior test performance compared to established ECG markers of AtCM and an alternative DL approach This also held on external validation sets. Importantly, the predicted imaging features improved risk stratification for HF in the UK Biobank, even when excluding patients with AF, suggesting that our model captures AtCM beyond AF.

**Discussion:** We established a novel DL approach for the diagnosis of AtCM from 12 lead ECG. Due to the wide availability of ECG, our approach has the potential to improve screening and diagnosis of AtCM.

**Structured Graphical Abstract:** *Key Question:* Can left atrial imaging markers of atrial cardiomyopathy predicted from 12 lead electrocardiogram (ECG) using a deep learning (DL) model improve risk stratification for atrial fibrillation (AF) and heart failure (HF)?

*Key Finding:* Our DL model successfully predicted left atrial imaging features from 12 lead ECG. The predicted left atrial imaging markers improved risk stratification for AF and HF outperforming a deep learning model trained to identify patients with AF directly.

*Take-home message:* DL allows improved diagnosis of atrial cardiomyopathy from ECG. Structured graphical abstract showing the model training and evaluation procedure. We first trained a deep learning model to predict left atrial imaging markers of atrial cardiomyopathy. We then added the predicted imaging features to clinical risk scores, creating multivariate regression models for risk stratification of different clinical outcomes. Finally, we externally validate the deep learning model and derived multivariate regression models on a diverse set of clinical cohorts.

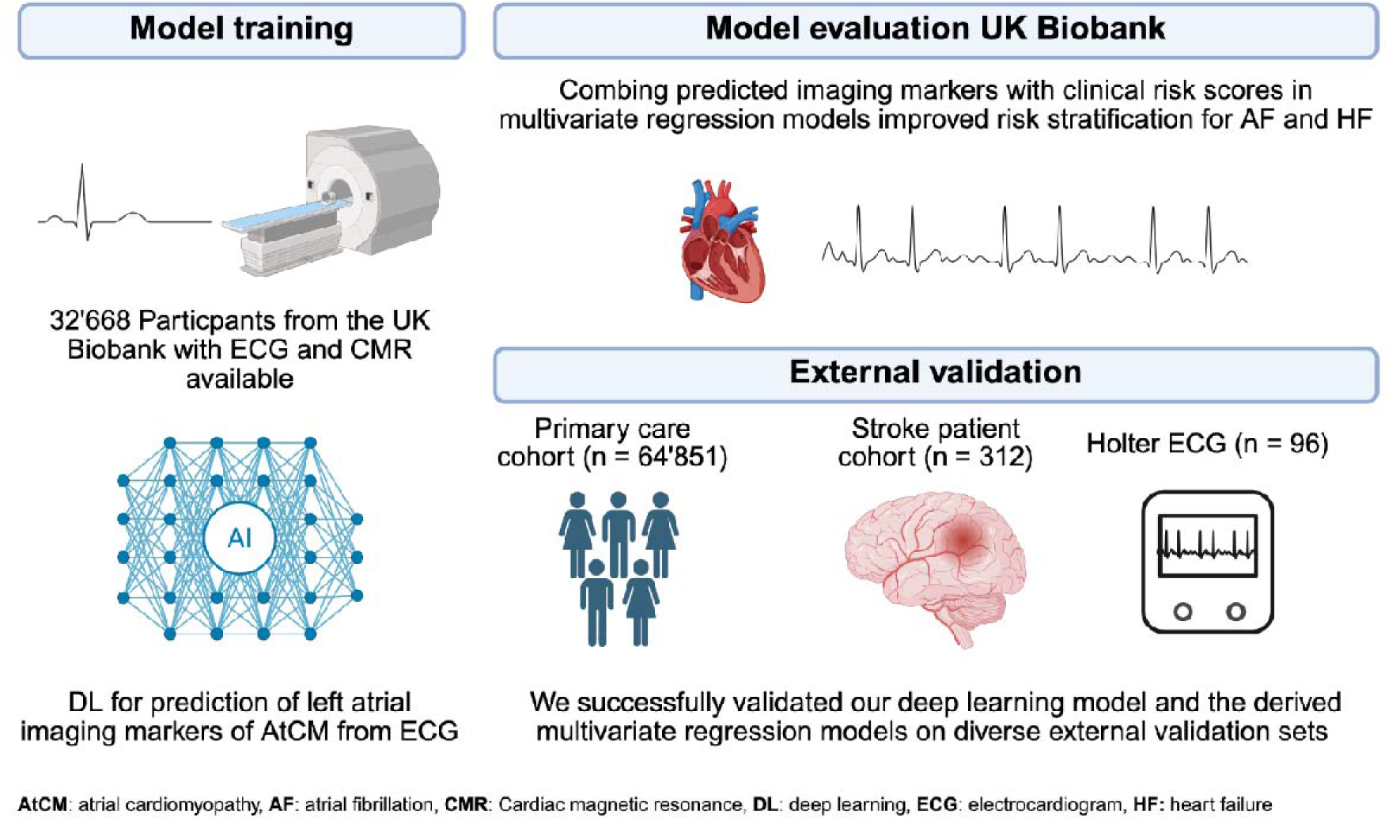

## 1. Introduction

Atrial cardiomyopathy (AtCM) is a condition arising from pathological structural and electrophysiological remodeling of the heart, particularly affecting the left atrium. Since AtCM potentially results in atrial fibrillation (AF), ischemic stroke and heart failure (HF) [1,2], early detection is essential. However, diagnosis of AtCM remains challenging [3]. Imaging markers including increased left atrial maximum volume (LA max) or left atrial minimum volume (LA min) indexed to body surface area (BSA), increased left atrial to left ventricular volume ratio (LALV) and reduced left atrial ejection fraction (LAEF) have been used to diagnose AtCM [3–7]. These imaging indices have been linked to AF, stroke and HF [8]. However, cardiac imaging is costly and not accessible in many regions. Blood biomarkers such as NTproBNP or MRproANP could aid in AtCM diagnosis, but they lack specificity and may fluctuate due to comorbidities or adjunctive drug therapies [1].

An electrocardiogram (ECG) based diagnosis of AtCM could address these limitations. ECG is a basic diagnostic tool in emergency departments and medical practices around the world. Specific ECG changes point towards the presence of AtCM, such as changes in P wave indices [9–13], but due to substantial variability, their use is not part of clinical routine [14].

Recently, the potential of artificial intelligence (AI) based analysis of ECG for diagnosis of AtCM has been recognized [1]. So far, in the field of ECG-based AI tools, deep learning (DL) approaches have focused on identifying patients with previous or incident AF [15–21], but not on the diagnosis of AtCM. However, the binary outcome (AF yes/no) does not capture the complex spectrum of changes that occur in AtCM, which include changes in atrial size and ejection fraction. This is reflected in current expert consensus, which highlights the importance of viewing AF and AtCM as separate entities [1]. Further, there is evidence that AtCM can contribute to adverse events such as HF or ischemic stroke even in the absence of AF. [1,2]. A DL approach with focus on the underlying structural and functional remodeling of the left atrium could capture more direct evidence, allowing a more accurate diagnosis of AtCM and an improved risk stratification for its sequelae. Further, it could facilitate the development of targeted treatments for AtCM, which have so far been hindered by lack of reliable diagnostic measures [1].

We here demonstrate development and validation of a new DL approach for diagnosing AtCM based on ECG. We finetuned an ECG foundational model (ECG-FM), pretrained on over 1 million ECG samples [22] to predict four left atrial imaging indices (indexed LA max, indexed LA min, LAEF and LALV) based on non-AF ECGs. We then leveraged the predicted left atrial imaging indices to build multivariate models to tackle different clinical challenges related to AF. An overview is shown in Figure 1. Further, we also validate our model on Holter-ECG with reduced leads. Finally, we aimed to investigate if our deep learning model could improve diagnosis and prediction of HF and whether it could do so independently of AF.

**Figure 1.**
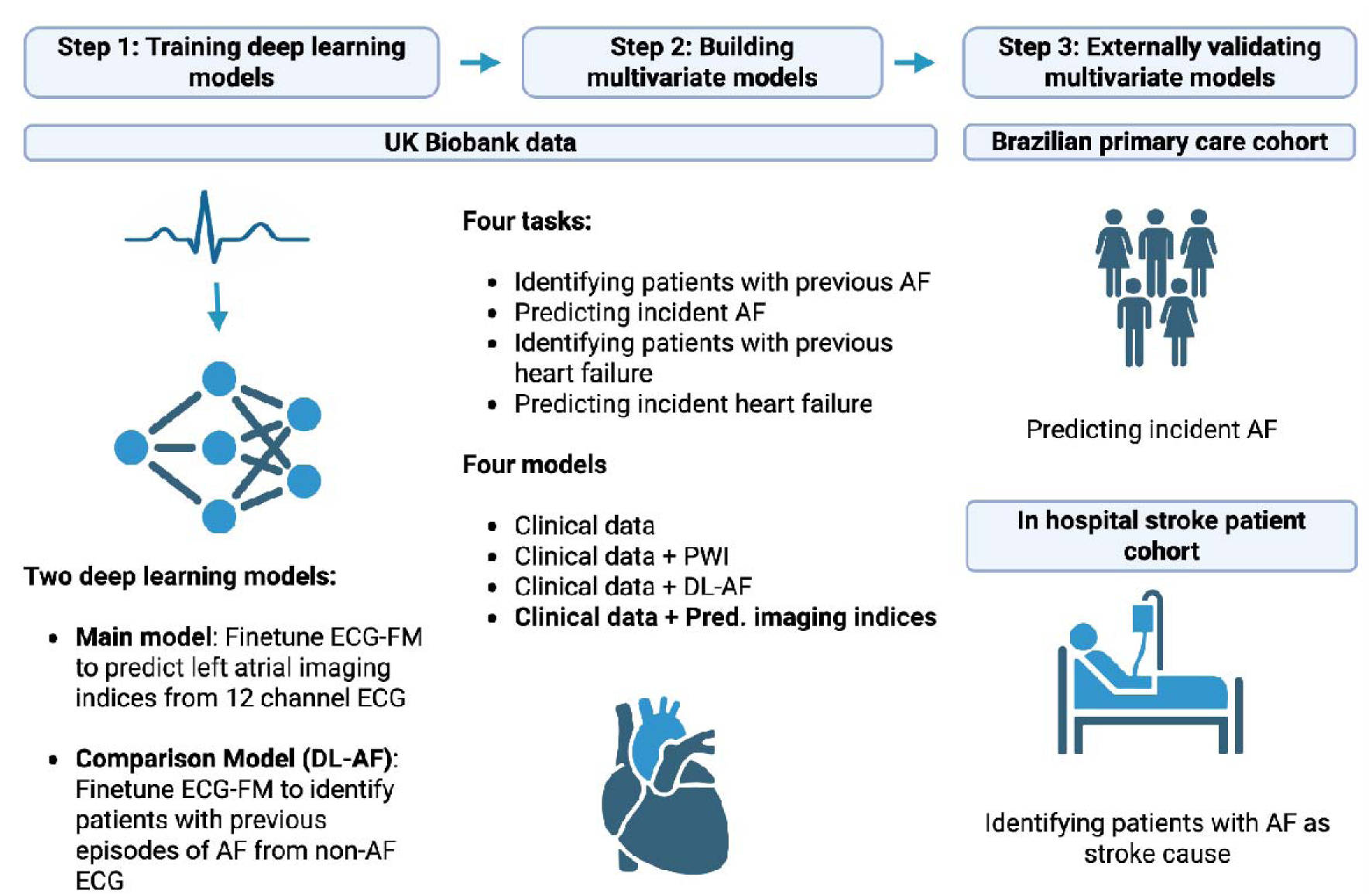
Workflow overview for model training and evaluation. AF: Atrial fibrillation. PWI: P wave indices.

## 2. Methods

### Data

Model training and evaluations were done on 12 lead ECGs. All ECGs which showed current AF were excluded. We performed preprocessing according to the specifications of McKeen et al. [22]. ECGs were resampled to 500 Hz using linear interpolation. As ECG-FM is based on 5 second 12 lead ECG segments, all ECGs were segmented into non-overlapping 12 lead 5 second segments. If ECGs were shorter than 10 seconds, the first 5 seconds were retained and the rest discarded. At test time, if two segments were available for a participant, the predictions of the model were averaged to obtain the final prediction. The datasets used are described in greater detail below.

### The UK Biobank (UKBB)

The UKBB is a large population-based cohort. The recruitment protocol is publicly available [23]. We worked with the data release from March 2025. As part of assessments, a portion of the participants underwent cardiac magnetic resonance (CMR) imaging and ECG according to standardized protocols [24,25]. From CMR imaging atrial and ventricular volumes have been previously segmented using deep learning and made available to researchers [26]. Details can be found in the supplemental methods.

Data of all participants with available imaging features and ECGs was divided into train, validation and test data using a 0.6/0.2/0.2 split. Both 5-second segments of each participant were allocated to the same split.

### Primary care cohort from Brazil

The Code15% dataset is a large, public ECG dataset consisting of 12 lead ECGs collected by the Telehealth Network of Minas Gerais between 2010 and 2016 and is comprised mainly of ECGs from primary care [27]. ECG diagnoses have been labelled by healthcare experts. All participants with multiple ECGs available were included. The earliest ECG of each included patient was taken as baseline and used for predicting left atrial imaging features. If the ECG showed AF, the patient was excluded. The dataset contains both 10 second and 7 second ECGs.

### Cohort of ischemic stroke patients from the USZ

Data of all ischemic stroke patients treated at the USZ in 2023 and 2024, who had not refused the use of their routine clinical data for research, were retrospectively collected. We received permission from the Cantonal Ethics committee Zurich to perform this data collection (PREDICT, BASEC-ID: PB_2016_01751 and PREDICT 2 BASEC-ID 2025-01797). All participants with at least one available non-AF ECG during in hospital stay and a determined stroke etiology were included in the analysis. The ECG with the highest quality control score as determined by a finetuned version of ECG-FM described in [22] was selected per patient.

In the USZ cohort, the downstream task was to identify patients where AF was identified as the cause of stroke by clinicians after complete diagnostic work up. This included both cases where AF was diagnosed before stroke, as well as cases where AF was identified during diagnostic workup of stroke etiology. Definition of stroke cause was done according to previous studies [28]. Details can be found in the supplemental methods.

### Holter ECG dataset

As all 12 leads of an ECG are not always available in clinical practice, we aimed to evaluate our model in a setting with reduced leads. The SHDB-AF dataset is a Holter ECG dataset containing 128 24-hour, 2 lead Holter ECGs as well as clinical data and left atrial diameter as measured by echocardiography. [29]. The two leads are NASA lead and CC5 lead. For 98 of the ECGs, beat level rhythm classifications are available. We selected these ECGs and extracted the first five minutes of consecutive non-AF recordings. We excluded all recordings without five minutes of consecutive non-AF recordings. We preprocessed these ECGs as described above, obtaining 60 5-second segments per ECG record. CC5 lead was taken as lead V5 and NASA lead as lead II [30]. The missing leads were masked with zeroes.

### Model Training and finetuning

We decided to finetune an ECG foundational model instead of training from scratch. Foundational models are trained on large datasets usually in a self-supervised way, thus reducing the need for labelled data. Numerous studies have shown that leveraging ECG foundational models improves results compared to training from scratch for different downstream tasks. [22, 31–33]. We used ECG-FM for our experiments [22]. Thus, we were able to leverage the more diverse, multicohort data including real world clinical datasets, that ECG-FM was pretrained on compared to training only on UKBB data. ECG-FM was pretrained on over 1 million training samples employing a hybrid self-supervised learning approach, which contained a contrastive learning task. To obtain positive pairs for contrastive learning, 10 second ECGs were segmented into two 5 second segments. Hence, the model requires 5 second segments as input. Importantly, neither UKBB data nor the Code15% dataset was used to pretrain ECG-FM.

We finetuned the model using multitask learning to predict indexed LA max, indexed LA min, LAEF and LALV from 12 lead ECG. We selected these four indices based on existing literature [3–8]. As AtCM is increasingly seen on a spectrum rather than a binary condition [1] we decided against combining the imaging indices with threshold to receive a binary label of AtCM. Details regarding finetuning can be found in the supplemental methods.

For model evaluation on the test set, we averaged the two predictions per patient (one for each 5 second segment). We plotted predicted versus ground truth values and calculated Pearson correlation coefficients as well as R^2^ values.

### Comparison models

We compared our novel method to two alternative approaches for diagnosing AtCM from 12 lead ECGs. The first is extracting four P wave indices (P wave duration, PR Interval, P wave amplitude and P Axis) which have been associated with atrial cardiopathy in literature [8–12]. Details can be found in the supplementary material. Second, as an alternative deep learning approach based on cross-sectional data, we finetune ECG-FM to identify patients with previous diagnosis of AF based on identical training and validation data and using identical training parameters. The only adjustment we made is to change the loss function from mean squared error loss to binary cross entropy loss, as it now is a classification rather than a regression task. We call this model deep learning AF (DL-AF). This approach reflects previous approaches to diagnose incident of previous AF from non-AF ECG using deep learning [15–21].

### Statistical analysis

All analysis of downstream tasks was done using R version 4.4.0. Using the predicted imaging indices, we tackled different clinically important tasks in the UKBB cohort. Only participants not included in train or validation set were included for fitting and evaluating multivariate models. We excluded participants with missing covariates. The first task was to identify participants with previous episodes of AF from non-AF ECG.

As complete clinical information is not always available, especially when performing population-based screening, we evaluated this task in two settings: one with limited clinical information comprised only of age and sex, the second with complete information on medical history and blood pressure measurements. For the limited setting we first built a logistic regression model using only age and sex, for the full information setting the base model contained all clinical parameters from the previously validated Cohorts for Heart and Aging Research in Genomic Epidemiology Atrial Fibrillation (CHARGE-AF) score [34]. Then we built three new logistic regression models by adding the four predicted imaging markers, the four P wave indices and the predicted probability of the DL-AF model respectively to the base model. We evaluated all four models using 10-fold cross validation. We calculate area under the receiver operating characteristic curve (ROC AUC) and area under the precision recall curve (PR AUC) as well as Brier scores with 95% confidence intervals for each of the models. We calculate P values for differences between ROC AUC values of the model with the predicted imaging features and the other features using DeLongs test [35]. Further, we perform sub-analyses stratified by sex. Finally, we trained the logistic regression models on the complete hold-out data from the UKBB and used the coefficients to calculate risk scores for external validation.

The second task in the UKBB was the prediction of five-year risk of incident AF. Participants with prior diagnosis of AF were excluded for this analysis. Instead of logistic regression models, cause specific hazard models were employed treating death as a competing event using the *survival* and *riskRegression* packages. Participants experiencing neither event until 01.01.2025 were censored. When evaluating predictive performance of the survival models we employed inverse probability of censoring weighting to account for censoring. ROC-AUC was calculated using the *riskRegression* package, PR-AUC was calculated using the *APtools* package. We also report Brier scores. We evaluated the models using 10-fold cross validation. Further, we perform a sub analysis stratified by sex. Again, we trained the cause specific hazard models on the complete hold-out data and used the coefficients to calculate risk scores for external validation.

Next, we investigated if the predicted imaging markers could also improve the diagnosis of HF beyond clinical data. We again started by building a multivariate logistic regression model based on clinical data. We based this model on the HF risk score proposed by Ho et al. [36] As data on subtype of HF is not available in the UKBB, we combined risk factors identified for prediction of HF with reduced ejection fraction and HF with preserved ejection fraction. Details can be found in the supplemental material. We then investigated if adding the predicted imaging markers improved detection of patients with previous diagnosis of HF, again comparing against the same alternative models, evaluating models using 10-fold cross validation.

As there is evidence that AtCM plays a role in the pathogenesis of HF independently of AF, we investigated how these results changed if patients with previously diagnosed AF were excluded. Further, we investigated if the predicted imaging markers improved the prediction of 5-year risk of incident HF beyond the HF risk score. We employed cause specific hazard models treating death as a competing event. We evaluated if the predicted imaging markers were associated with incident HF when adjusting for the parameters in the HF risk score [36]. Then we compared predictive performance of the different models using 10-fold cross validation. Again, we investigated how the results changed when removing patients with previous diagnosis of AF and censoring patients with incident AF diagnosis.

We externally validated the multivariate model for predicting incident AF in the Brazilian primary care cohort. We used the regression coefficients of the multivariate models for prediction of incident AF, containing age and sex as clinical parameters from the UK Biobank. No other clinical parameters are available in this cohort. For calculation of brier scores, as the dates of the follow up ECGs and thus length of follow-up are not known in this cohort, we recalibrated the model so that the average prediction of the models matched the prevalence of incident AF in the cohort. We performed sub analyses stratified by sex and by age, comparing patients at ages 65 or lower and above 65, as opportunistic AF screening with 12 lead ECG has been previously recommended especially in patients above 65 years [37].

In the cohort of ischemic stroke patients, the downstream task was to identify patients in which AF was diagnosed as stroke cause. We evaluated both the models including only age and sex and including the full clinical information (excluding race, as this was not available) for detecting patients with previous diagnosis of AF. Details on definition of covariates can be found in the supplementary material.

In the SHDB-AF dataset we averaged the predictions per recording over all 60 samples. We then investigated if the average predicted indexed LA maximum volumes and predicted LA minimum volumes correlated with the left atrial diameter measured in echocardiography. Left atrial diameter and left atrial volume have been shown to be correlated [38]

### Model explainability

To visualize which parts of the ECG were important for prediction of the four imaging markers, we employed vanilla gradients [39] calculating saliency maps for each of the four predicted outputs based on the ECGs from the University Hospital Zurich ischemic stroke patient cohort. To quantify the results, ECGs were segmented into P wave, PQ interval, QRS complex, ST segment, T wave and end of T wave to next P wave onset using neurokit2. Details can be found in the supplementary methods. Absolute saliency values were averaged for each segment, then averaged over all 12 leads and finally divided by total average saliency for each ECG. Boxplots were used to show the results. To validate our findings, we repeated the analysis using integrated gradients.

## 3. Results

### Predicting left atrial imaging indices from routine 12 lead ECG

First, to predict atrial imaging indices from routine 12 lead ECG, we finetuned the ECG-FM model [22] on 19’600 ECGs from the UK Biobank (UKBB) cohort in the train set, 6’534 ECGs in the validation set and 6’534 ECGs in the test set. An overview of inclusion and exclusion criteria for the data used can be found in Figure 2. The finetuned model’s predictions of atrial imaging indices showed highly significant (p < 0.001) correlations with the ground truths from cardiac imaging, ranging from 0.41 – 0.52. (Figure 3)

**Figure 2.**
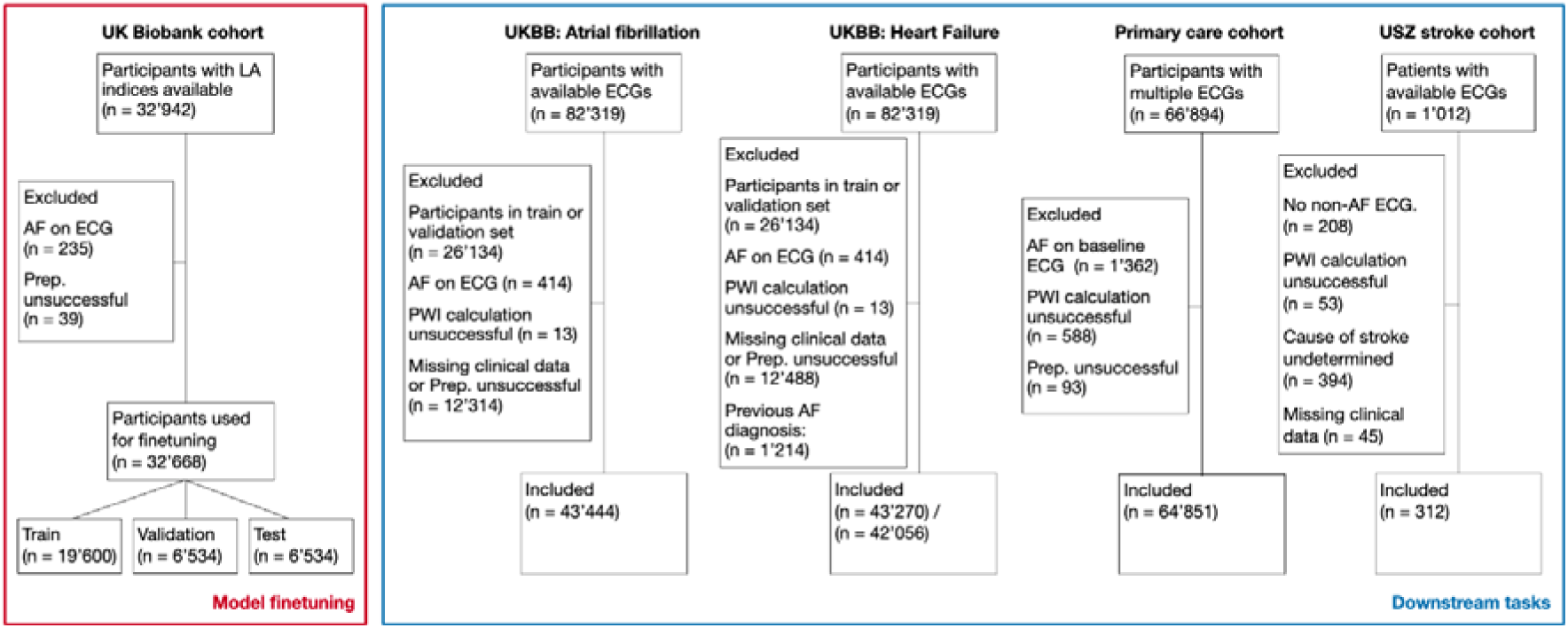
Overview over participant inclusion and exclusion for model finetuning (left) and clinical tasks (right). In UKBB: Heart Failure the final numbers represent included participants with and without exclusion of patients with previous AF. UKBB: UK Biobank, LA: left atrial, Prep: preprocessing, AF: atrial fibrillation, PWI: P wave indices, ECG: electrocardiogram.

**Figure 3.**
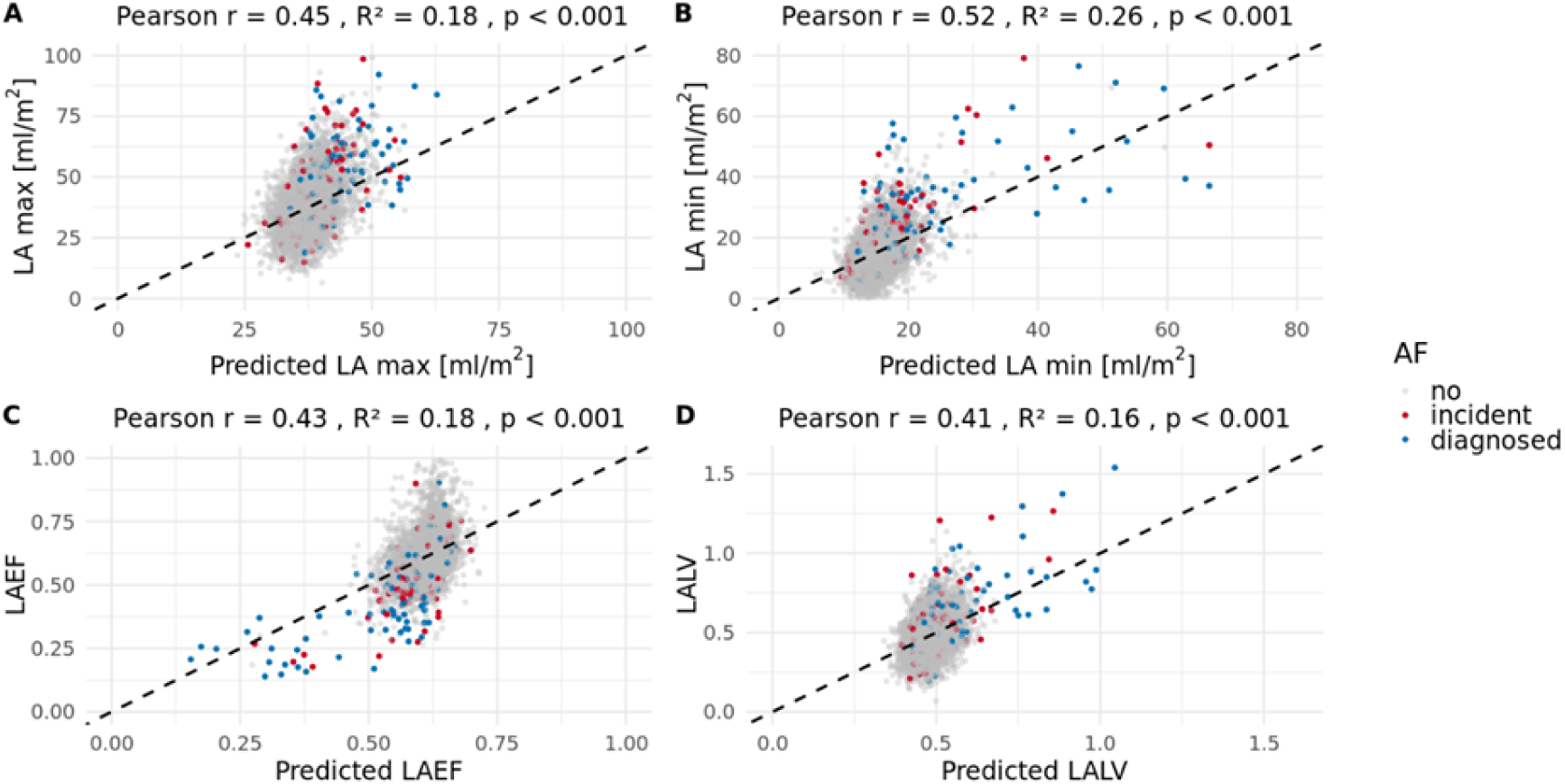
Scatter plot of ground truth and test set predictions for atrial imaging indices: indexed left atrium maximum volume (A), indexed left atrium minimum volume (B), left atrium ejection fraction (C) and left atrial to ventricular volume ratio (D). Dotted lines show perfect predictions. Colors indicate if atrial fibrillation had been diagnosed and documented in health records before the imaging date (blue) or if incident atrial fibrillation occurred at some point after imaging (red). Grey points indicate participants without any documented atrial fibrillation. LA max: indexed left atrial maximum volume, LA min: indexed left atrial minimum volume, LAEF: left atrial ejection fraction, LALV: left atrial to ventricular volume ratio.

### Identifying patients with previous episodes of AF and incident AF in the UKBB

To apply the predicted atrial imaging indices to the prediction of previous and incident AF, we trained and evaluated multivariate regression models, based on the predictions of the deep learning models and clinical parameters. We trained these multivariate models on data from the UKBB. Population characteristics can be found in Table 1, inclusion and exclusion criteria in Figure 2. Results of multivariate models for identifying participants with previous AF and predicting incident AF are shown in Table 2 and Figure 4. Adding the four predicted imaging markers to the clinical parameters from the CHARGE-AF score [33], significantly improved both detection of patients with previous episodes of AF from non-AF ECG (ROC-AUC 0.756 (0.742 – 0.771) vs. 0.720 (0.706 – 0.735), p < 0.001) and 5-year risk prediction of incident AF (ROC-AUC: 0.777 (0.747 – 0.807) vs. 0.758 (0.728 – 0.788), p = 0.018). Further, to compare our novel approach to previously reported approaches of diagnosing AtCM from ECG, we trained two comparison models on the UKBB data. The first is based on traditional P wave indices extracted from 12 lead ECG which have been previously linked to AtCM [8–12], the other is an alternative deep learning approach which reflects previous approaches reported in literature (DL-AF) [15–21]. Our novel approach of predicting left atrial indices, outperformed both alternative approaches (Table 2, Figure 4). When only using limited clinical parameters (age + sex) the model including predicted imaging indices also outperformed all comparison models (Table S1). Regression coefficients of the multivariate models can be found in the Supplementary Data Table 1. The models containing the predicted imaging indices outperformed in other models in both sexes. (Table S2-S5)

**Figure 4.**
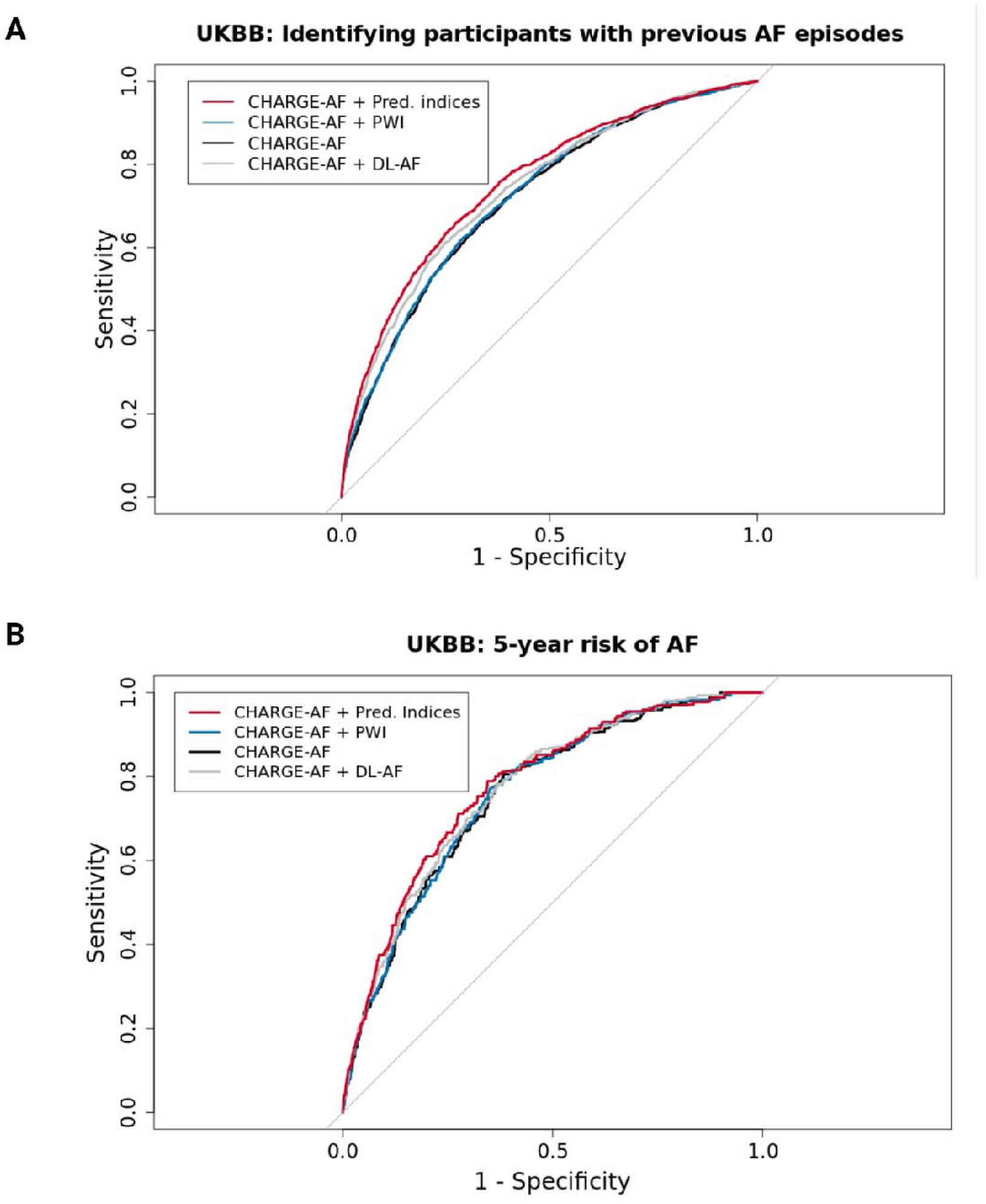
Receiver operating characteristic curves for identifying patients with previous episodes of atrial fibrillation (A) and for predicting 5-year risk of new onset atrial fibrillation (B) in the UK Biobank cohort. AF: atrial fibrillation, pred. indices: predicted imaging indices, PWI: P wave indices, DL: deep learning. UKBB; UK Biobank.

**Table 1.**
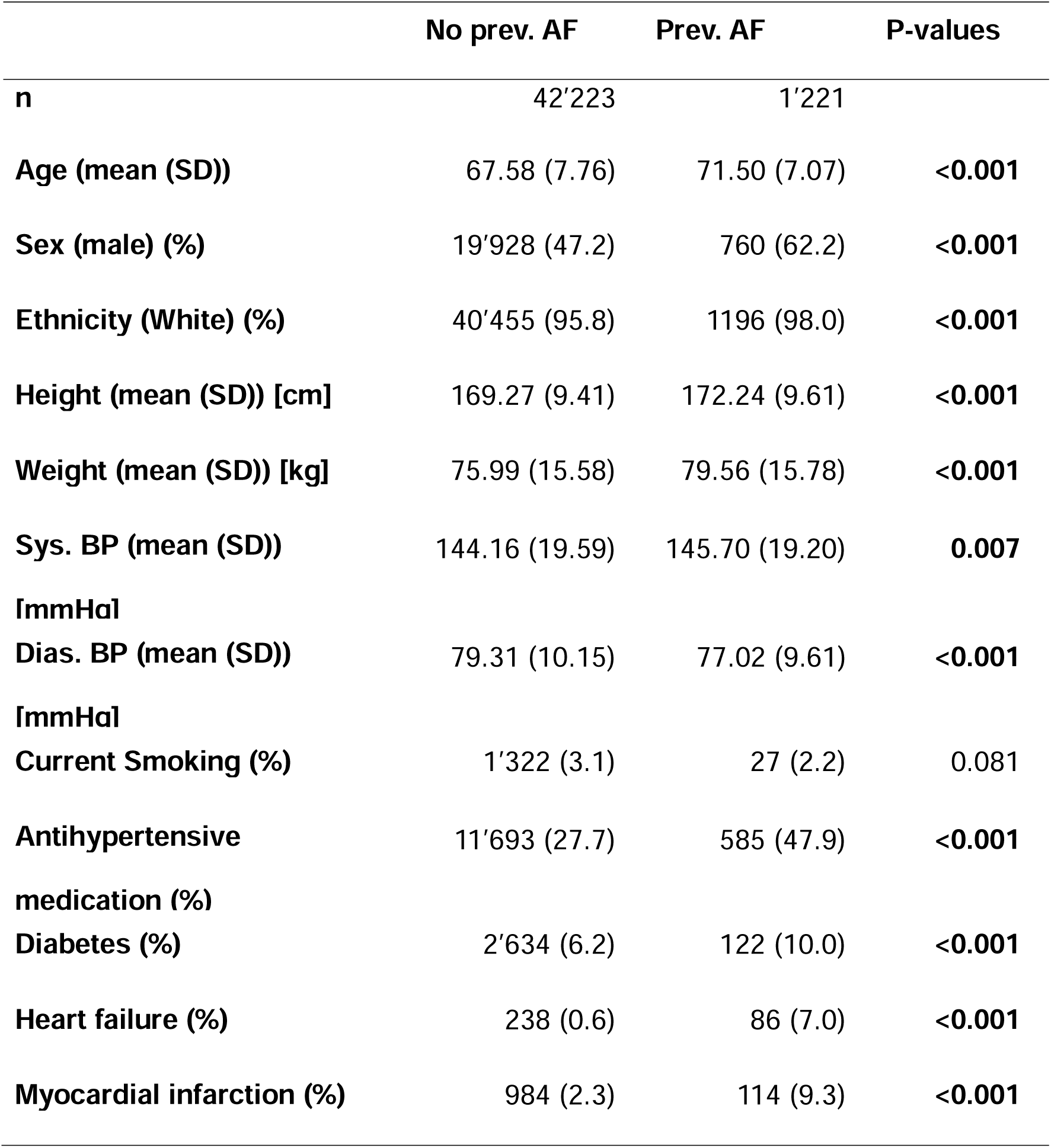
Participant characteristics in the UK Biobank cohort used for evaluation of downstream tasks. Stratification was done according to previous episodes of atrial fibrillation documented in health records. Prev: Previous, SD: standard deviation, sys. BP: systolic blood pressure, dias. BP: diastolic blood pressure. For age, height, weight, systolic blood pressure and diastolic blood pressure p-values were calculated using two-tailed unpaired T-test, for the other variables Pearsons chi-squared test was used.

**Table 2.** ROC-AUC and PR-AUC values with 95 % confidence intervals for model performances on the different downstream tasks in internal and external validation. Our new proposed method is shown in bold. AF: Atrial fibrillation, Pred. indices: predicted imaging indices. PWI: P wave indices, DL: deep learning, ROC-AUC area under the receiver operating characteristic curve, PR-AUC: area under the precision recall curve. UKBB: UK Biobank, USZ: University Hospital Zurich. 95% confidence intervals were calculated using 1000 bootstrap samples. Brier score in %. Maximum is 100% minimum is 0%. Lower values indicate better calibration. P-values were calculated by comparing ROC-AUC using DeLongs test. We performed pairwise comparisons between the model performance of the model containing the predicted imaging indices and the performance of the other models.

| UKBB: Identifying participants with previous AF episodes |  |  |  |  |
| --- | --- | --- | --- | --- |
| Model | ROC – AUC | PR-AUC | Brier (%) | P - values |
| **CHARGE-AF + Pred. indices** | 0.756 (0.742-0.771) | 0.115 (0.100-0.130) | 2.62 (2.48-2.76) |  |
| CHARGE-AF + PWI | 0.724 (0.710-0.738) | 0.090 (0.077-0.102) | 2.66 (2.51-2.79) | **< 0.001** |
| CHARGE-AF | 0.720 (0.706-0.734) | 0.087 (0.075-0.099) | 2.66 (2.51-2.80) | **< 0.001** |
| CHARGE-AF + DL-AF | 0.741 (0.727-0.755) | 0.106 (0.092-0.120) | 2.64 (2.45-2.78) | **< 0.001** |
| N = 43'444, AF = 1'221 Prevalence: 0.028 |  |  |  |  |
| UKBB: 5-year risk of AF |  |  |  |  |
| Model | ROC-AUC | PR-AUC | Brier (%) | P – values |
| **CHARGE-AF + Pred. indices** | 0.777 (0.747-0.807) | 0.033 (0.026-0.047) | 1.28 (1.11-1.45) |  |
| CHARGE-AF + PWI | 0.760 (0.731-0.790) | 0.025 (0.020-0.031) | 1.29 (1.12-1.46) | **0.046** |
| CHARGE-AF | 0.758 (0.728-0.788) | 0.025 (0.020-0.031) | 1.29 (1.12-1.46) | **0.018** |
| CHARGE-AF + DL-AF | 0.770 (0.740-0.799) | 0.029 (0.023-0.041) | 1.28 (1.11-1.45) | 0.27 |
| N = 42'223, AF = 248 Event rate = 0.013 |  |  |  |  |
| Primary care cohort: Predicting future AF |  |  |  |  |
| Model | ROC-AUC | PR-AUC | Brier (%) <sup>†</sup> | P – values |
| **Age + Sex + Pred. indices** | 0.849 (0.838-0.860) | 0.220 (0.195-0.245) | 1.68 (1.59–1.77) |  |
| Age + Sex + PWI | 0.784 (0.773-0.796) | 0.058 (0.053-0.064) | 1.86(1.76- 1.95) | **< 0.001** |
| Age + Sex | 0.771 (0.760-0.782) | 0.051 (0.046-0.055) | 1.86 (1.77–1.96) | **< 0.001** |
| Age + Sex + DL-AF | 0.829 (0.818-0.840) | 0.183 (0.160-0.207) | 1.73 (1.65–1.83) | **< 0.001** |
| N = 64'851, AF = 1'255 Prevalence: 0.019 |  |  |  |  |
| USZ: Diagnosing patients with AF as cause of stroke |  |  |  |  |
| Model | ROC-AUC | PR-AUC | Brier (%) | P – values |
| **CHARGE-AF + Pred. Indices** | 0.767 (0.706-0.828) | 0.429 (0.315-0.544) | 1.81 (1.42-2.22) |  |
| CHARGE-AF + PWI | 0.741 (0.681-0.800) | 0.395 (0.287-0.504) | 1.82 (1.44–2.21) | 0.14 |
| CHARGE-AF | 0.732 (0.669-0.795) | 0.390 (0.281-0.498) | 1.85 (1.46-2.26) | **0.036** |
| CHARGE-AF + DL | 0.745 (0.687-0.803) | 0.415 (0.297-0.533) | 1.81 (1.43-2-18) | 0.12 |
| N = 312, AF = 65 Prevalence = 0.208 |  |  |  |  |

### Identifying patients with previous diagnosis of HF and incident HF in the UKBB

Since AtCM also plays a role in HF, we investigated whether, similar to our AF analysis, our DL approach could enhance the detection of patients with a prior HF diagnosis and improve the prediction of incident HF. Patient characteristics can be found in Table S6. The model containing the predicted imaging markers significantly outperformed the comparison models for detection of patients with previous diagnosis of HF. (Table S7). When removing all patients with previous diagnosis of AF, the model containing the predicted imaging markers still outperformed the other models, although differences only partially reached statistical significance, due to the lower number of cases after removing participants with AF (Table S8). Importantly the model containing the predicted imaging indices was the only one to consistently outperform the HF risk score across all metrics when removing patients with AF.

Next, we investigated associations of the predicted imaging indices with incident HF. All four predicted imaging indices were significantly associated with incident HF when adjusting for the risk factors from the HF risk score (Table S9). Although the size of these associations decreased, when censoring patients with AF diagnosis, they remained significant for predicted LA max, LA min and LAEF (Table S10). While LALV still showed an association with incident HF, this lost significance. (Table S10). Further, adding the predicted imaging markers to the HF risk score led to modest improvements in ROC-AUC and PR-AUC, which remained when censoring patients with AF. However, these differences did not reach statistical significance. (Tables S7, S8)

### External validation in a primary care cohort

While the UKBB is a valuable resource, its highly standardized protocol does not necessarily reflect real world clinical practice. Hence, we aimed to validate our model in real world healthcare cohorts. First, we validated our model in the Code15% dataset, comprised of ECGs collected mainly during primary care assessments in Brazil [26]. 64’851 of the patients in the cohort had a follow-up ECG. Patients in the primary care cohort were younger on average (59 years ± 19 years) with a larger age range represented than in the UKBB. Also, patients were predominantly female (62%). In the primary care cohort, the model containing the predicted imaging indices significantly outperformed the three comparison models for identifying patients with incident AF (p < 0.001) (Table 2 and Figure 5A). The model containing the predicted imaging indices outperformed the other models in both sexes and both evaluated age groups. (Tables S11-S12)

**Figure 5.**
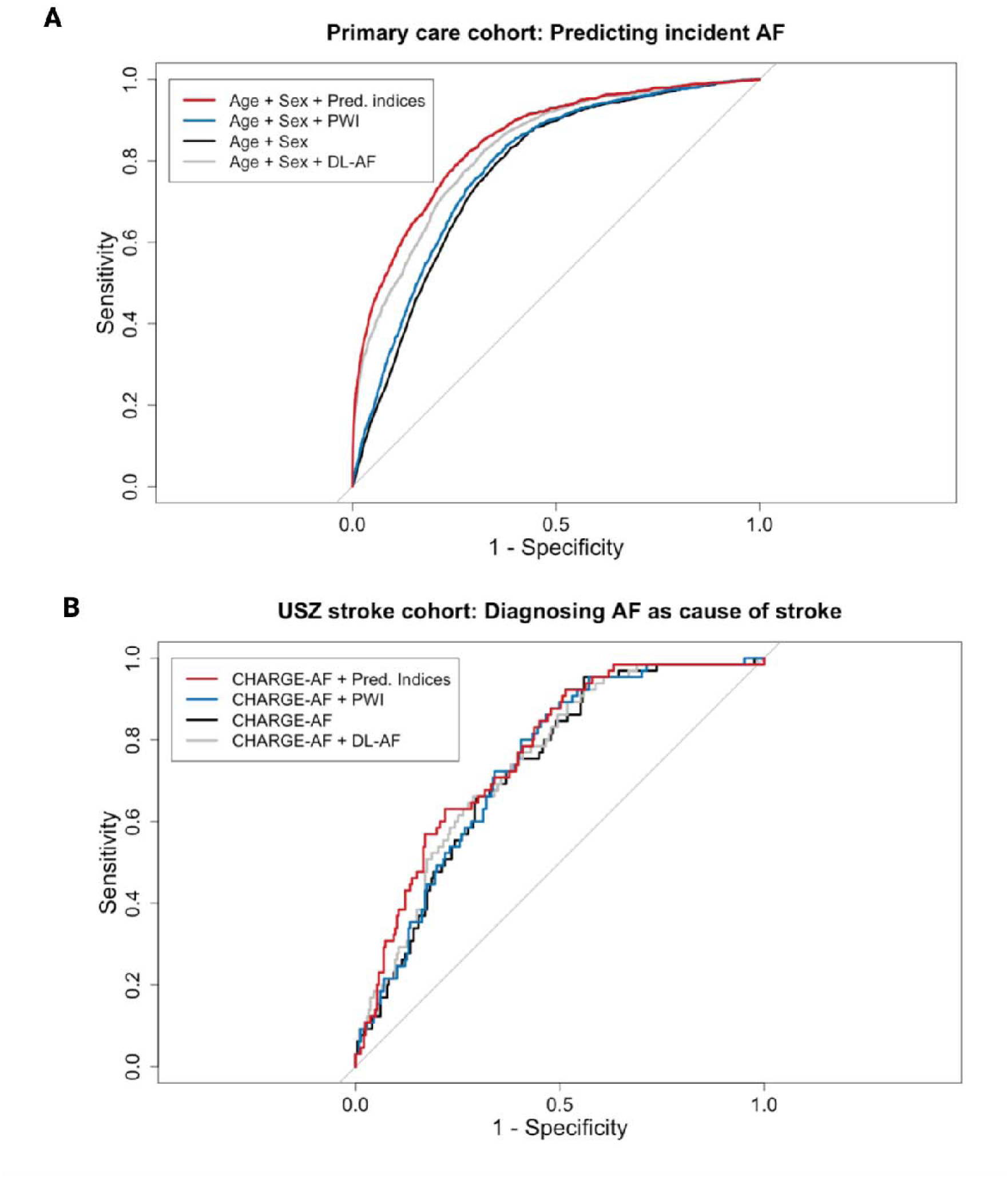
External validation receiver operating characteristic curves for identifying patients with incident AF in a primary care cohort (A) and for identifying patients with atrial fibrillation as cause of stroke in an in-hospital cohort of ischemic stroke patients (B). AF: atrial fibrillation, pred. indices: predicted imaging indices, PWI: P wave indices, DL: deep learning.

### External validation in a cohort of ischemic stroke patients

In a next step, we validated our multivariate models in a cohort of ischemic stroke patients from the University Hospital Zurich (USZ), aiming to identify stroke patients with AF as cause of stroke. 312 patients (mean age 64 ± 17 years; 61% male) treated for ischemic stroke at the USZ in 2023 and 2024 with identified cause of ischemic stroke were included for the model evaluation. Results when combining the predicted imaging features with the CHARGE-AF score (Table 2, Figure 5B) and age and sex (Table S1) were similar, with the model containing the predicted imaging indices consistently outperforming the comparison models.

### Analyzing Holter ECG with reduced leads

While 12 lead ECG is most widely performed in clinical practice, there are numerous scenarios in which only a reduced number of leads is available, for example in long term rhythm monitoring. Hence, we evaluated the applicability of the model for analyzing Holter ECG with only two instead of 12 leads. We analyzed data from the Saitama Heart Database Atrial Fibrillation (SHDB-AF) dataset [29]. In the SHDB-AF dataset, two recordings were removed due to no 5-minute interval of consecutive non-AF ECG being available, leaving 96 recordings to analyze. Average age was 68 ± 11 years and 54% of patients were male. Both average, predicted indexed LA max (r = 0.47, 95% CI 0.29 - 0.61, p < 0.001) and predicted indexed LA min (r = 0.42, 95% CI 0.23 – 0.57, p < 0.001) were highly significantly correlated with the left atrial diameter measured in echocardiography as would be expected for left atrial volumes [38].

### Model Explainability

We then analyzed which parts of the ECG were important for the model’s predictions of atrial imaging indices using vanilla gradients [39], based on the 5 second segments of the ECGs from the USZ stroke patients. Looking at the saliency maps, the P wave morphology was important for predictions. An example is shown in Figure 6F. To quantify this, we segmented ECGs into P wave, PQ interval (end P wave to Q peak), QRS complex (Q peak to S peak), ST segment (S peak to T wave onset) and TP interval (T wave offset to next p wave onset). A schematic is shown in Figure 6E. We then calculated mean absolute saliency for each segment per lead, divided by mean saliency of the whole 5 second ECG segment per lead. We then averaged results from all 12 leads. This confirmed our qualitative findings as mean relative saliency was highest for the P wave segment. (Figure 6). Using integrated gradients produced similar results (Figure S1).

**Figure 6.**
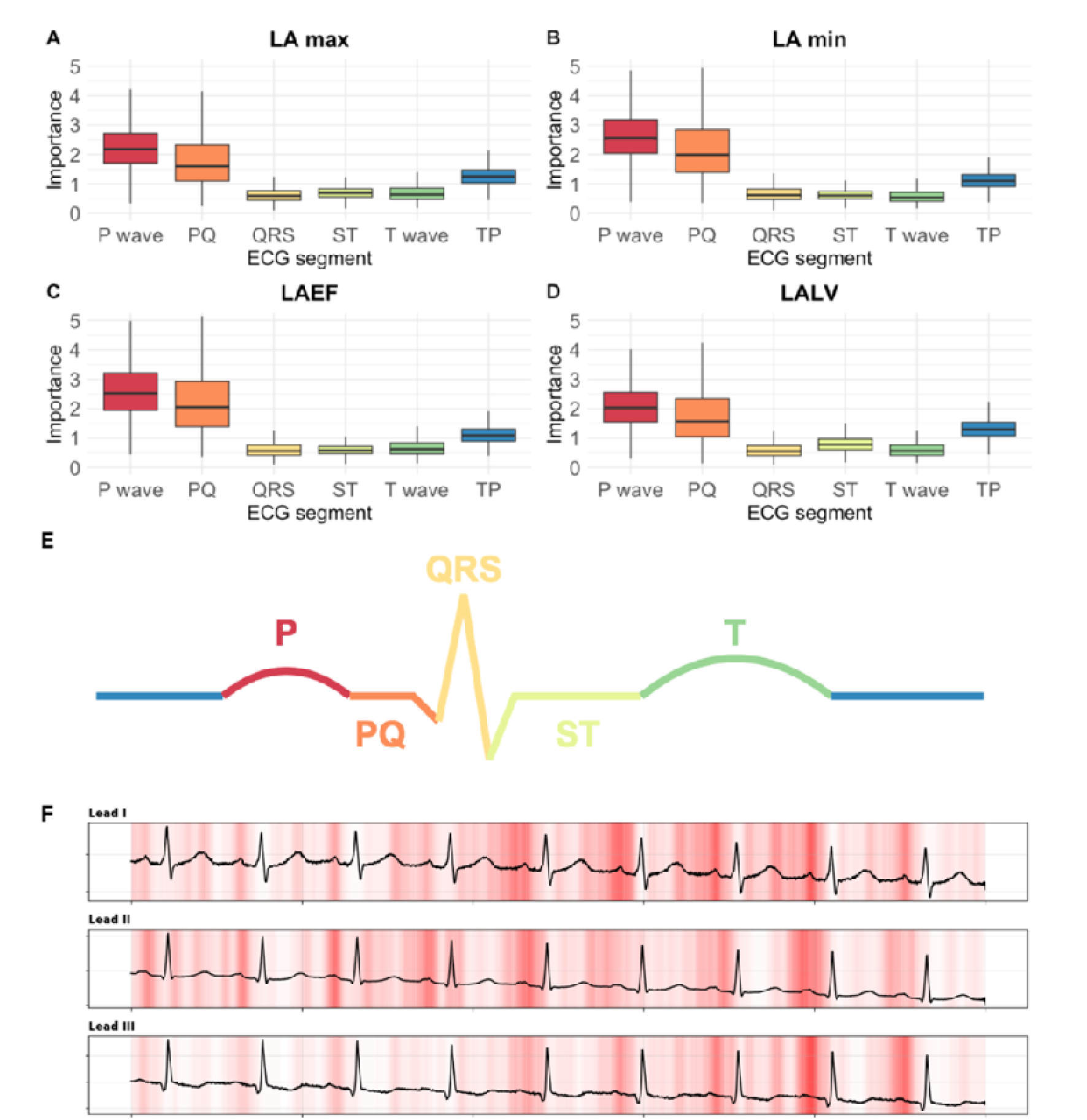
Importance of different ECG segments calculated using vanilla gradients for prediction of indexed left atrial maximum volume (A), indexed left atrial minimum volume (B), left atrial ejection fraction (C) and left atrial to ventricular volume ratio (D) evaluated on 1’077 5 second non-AF ECG samples from 653 patients from the USZ stroke cohort over all 12 leads. Importance was calculated by averaging absolute saliency values over all 12 leads per segment and then dividing by average salience of the entire 5 second ECG sample. (E) Schematic representaion of the ECG segments evaluated in (A)-(D). (F) Example saliency map for leads I-III. Darker red indicates high absolute saliency. LA max: indexed left atrial maximum volume, LA min: indexed left atrial minimum volume, LAEF: left atrial ejection fraction, LALV: left atrial to ventricular volume ratio.

## 4. Discussion

Our study demonstrates that left atrial CMR indices indicative of AtCM can be predicted from routine 12 lead non-AF ECG. The predicted imaging indices improved established models for tackling important clinical tasks, including the identification of participants with previous and incident AF and HF, and the detection of ischemic stroke patients with AF as underlying cause of stroke, based on a short, standard non-AF ECG. Hence, we provide a promising new approach for diagnosing AtCM from 12 lead ECG in patients who require timely detection and who may benefit from adjustment in preventive treatments.

The multivariate regression models we propose and validated for detection of AF address important clinical needs. Identifying patients at risk of developing future AF could lead to more stringent controls and better prevention of AF or AF-related complications [40]. For primary prevention of ischemic stroke, detecting patients with previous episodes of AF is important, as AF is associated with an approximately five-fold increase in risk of ischemic stroke, which can be reduced through targeted prophylaxis with anticoagulation. [41,42]. However, detecting patients with previous episodes of AF is challenging in clinical practice as it is commonly asymptomatic and intermittent, requiring long-term rhythm monitoring [43]. In patients with ischemic stroke with AF as the underlying etiology, anticoagulation is even more important, as it reduces the risk of stroke recurrence by more than half [44]. Hence, our presented multivariate models can be valuable tools for 1) preventing AF and 2) preventing ischemic stroke.

We further demonstrate that our model also provides meaningful predictions when analyzing Holter ECG, when only two leads are available, different to the ones the model was trained on. This highlights the versatility of our model and broadens the range of potential applications.

Further, there is increasing evidence that AtCM plays a role in stroke risk and HF independently of AF [1]. We show that our new approach enhances detection of patients with previously diagnosed HF and prediction of incident HF even when removing patients with known AF and censoring patients with incident AF. This contrasts the other evaluated approaches for diagnosing AtCM from ECG, which did not consistently improve predictions over the clinical risk score. These results show the promise of our DL- approach to capture AtCM beyond AF.

Prior studies have analyzed ECG using deep learning [15–21]. However, these studies have focused on identifying participants with previous or incident AF, not on diagnosing AtCM. To compare our approach to these previously described ones, we finetuned the ECG-FM model [22] to identify patients with previous episodes of AF directly, based on 12 lead ECG using identical training and test data. Our newly proposed approach based on predicting atrial imaging indices consistently outperformed this comparison model, across different cohorts and tasks. (Table 2) Further we provide first evidence that our approach could diagnose AtCM beyond AF by showing it allowed detection and prediction of HF even in patients free of AF, which contrasts the model trained to detect AF directly.

There are several potential reasons why prediction of atrial imaging indices may be superior to prediction of AF when attempting to diagnose AtCM. Firstly, there is increasing evidence that AtCM can develop (accompanied by changes in ECG and cardiac imaging) even in the absence of AF [1]. Secondly, a binary classification of AF does not adequately reflect the different severities of AF from short-lasting paroxysmal AF to long lasting permanent AF. These limitations can lead to bias in model training. Atrial imaging markers on the other hand, can identify AtCM even if it does not manifest as AF [1] and correlate more strongly with more severe, long-lasting forms of AF [45,46]. Hence, our approach better captures the full spectrum of AtCM, leading to an improved risk stratification.

The fact that the P wave segment was the most important ECG segment for model prediction makes sense, as the P wave represents the depolarization and conduction of the electric impulse from the atria to the ventricle. Thus, saliency maps allowed insights into variables important for model classification, increasing confidence in our model.

### Strengths and Limitations

The extensive external validation of our model over multiple real-world clinical datasets is a strength of our study. Importantly, we also validated our model on two non-European cohorts, a primary care cohort from Brazil and the Japanese SHDB-AF dataset. Finally, working with an ECG foundational model allowed us to leverage large amounts of unlabeled ECG data. Performing full finetuning and employing multilayer regression heads ensured the necessary flexibility to adapt to the new task.

A limitation of our study is that in the UKBB and in the primary care cohort no systematic long-term rhythm monitoring was undertaken. Therefore, some participants might have had undiagnosed, paroxysmal AF. Further, the UKBB is a population-based cohort and hence showed low prevalence of AtCM and AF. We decided against up sampling due to lack of clear criteria for selecting positive cases with AtCM. Training our model on datasets with higher prevalence of AtCM might further improve results. Further, additional imaging parameters such as left atrial strain are increasingly employed to diagnose AtCM [47]. Predicting left atrial strain from ECG could further improve diagnosis of AtCM. However, atrial strain was not measured as part of the UKBB imaging protocol. The external validation in the Brazilian primary care cohort also has its limitations, as only age and sex were available for this cohort. Hence the improvements in model performance when adding the predicted imaging features might partially be due to confounding cardiovascular risk factors. Finally, our analysis concerning HF as an endpoint is limited by the low number of patients diagnosed with HF in the UKBB, leading to only partial significance of results.

### Outlook

There are important considerations to be made before our model can be implemented in clinical practice. As the predicted imaging markers only showed moderate correlation with ground truth imaging indices, they cannot be interpreted as if they were actual imaging markers. Instead, our multivariate regression models should be employed. The regression models should be validated as screening tools in prospective clinical trials. For example, AF risk could be determined in patients free of prior AF diagnosis using a baseline ECG. Then, study participants could be asked to undergo rhythm monitoring, to investigate if patients with high predicted AF risk, show higher rates of incident AF. Similar studies have been proposed for validation of non-ECG based AF risk scores [48].

### Conclusions

We describe a novel DL approach for diagnosing AtCM from ECG and demonstrate the utility of the derived multivariate regression models across multiple cohorts and clinical tasks. Predicting left atrial imaging indices rather than clinical endpoints such as AF directly, improved risk stratification for AF across different cohorts. Further, it allows the diagnosis of underlying AtCM whose diagnosis is difficult but needed for the development and administration of targeted treatments [1]. Importantly, we provide first evidence that our approach has the potential to diagnosis AtCM beyond AF. Our findings highlight the potential of an AtCM-centered DL approach for better prevention of AF, HF and stroke.

## 5. Acknowledgements

This research has been conducted using the UK Biobank Resource under application number 467527. This work uses data provided by patients and collected by the NHS as part of their care and support. Figure 1 was created using BioRender.

## 6. Funding

This work was supported by the Swiss National Science Foundation (SNSF 310030_200703) and the UZH Candoc Grant (25-024).

## 7. Disclosures of interest

JS has received consultant and / or speaker fees from Abbott, Bayer, Berlin-Chemie, Biosense Webster, Biotronik, Boehringer-Ingelheim, Boston Scientific, Daiichi Sankyo, Medscape, Medtronic, Menarini, Pfizer, Saja, and WebMD. He reports ownership of Swiss EP and CorXL.

AB has received consulting/presenter fees from Abbott, Angiodynamics, Bayer Health Care, Biosense Webster, Biotronik, BMS/Pfizer, Boston Scientific, Cook Medical, Daiichi Sankyo, Medtronic, Philips, and Zoll.

SW received research funds by the Swiss National Science Foundation, the UZH Clinical research priority program (CRPP) stroke, the Zurich Neuroscience Center (ZNZ), the Baugarten foundation, the Hartmann Müller Foundation, the Koetser Foundation, the Philas Foundation, the Swiss Heart Foundation; and speaker honoraria from Amgen, Springer, Advisis AG, Teva Pharma, Boehringer Ingelheim, Lundbeck, Astra Zeneca, FoMF, and a consultancy fee from Bayer and Novartis via institution for research.

The remaining authors report no conflicts of interest.

## 8. Data availability statement

UK Biobank will make the data available to all bona fide researchers for all types of health-related research that is in the public interest. For more details on the access procedure, see the UK Biobank website: http://www.ukbiobank.ac.uk/register-apply/. Data from the Code15% dataset is publicly available under https://doi.org/10.5281/zenodo.4916205. The SHDB-AF dataset is publicly available under https://doi.org/10.13026/f5mp-gx65. The clinical data analyzed in this study will be made available upon reasonable request

The code for the analysis is available under: https://github.com/jul-des/DL-AtCM.git

The finetuned model weights will be made available via the UK Biobank, in accordance with its rules and regulations.

## 9. Supplemental Material

Detailed Methods

Tables S1-S16

Figure S1

Supplemental Data Table 1

## Notes

### Funding Statement

This work was supported by the UK Biobank Platform Credits Programme for covering computational costs, the Swiss National Science Foundation (SNSF 310030_200703) and the UZH Candoc Grant (25-024).

### Author Declarations

Kantonal ethics committee Zurich

### Summary of Updates

Added analyses on predicting heart failure, as well as age and sex stratified analyses. Added outlook on clinical implementation to discussion.

## References

1. Goette A, Corradi D, Dobrev D, Aguinaga L, Cabrera JA, Chugh SS, et al. Atrial cardiomyopathy revisited-evolution of a concept: a clinical consensus statement of the European Heart Rhythm Association (EHRA) of the ESC, the Heart Rhythm Society (HRS), the Asian Pacific Heart Rhythm Society (APHRS), and the Latin American Heart Rhythm Society (LAHRS). Europace. 2024 Aug 30;26(9). DOI: 10.1093/europace/euae204

2. Edwards JD, Healey JS, Fang J, Yip K, Gladstone DJ. Atrial Cardiopathy in the Absence of Atrial Fibrillation Increases Risk of Ischemic Stroke, Incident Atrial Fibrillation, and Mortality and Improves Stroke Risk Prediction. J Am Heart Assoc. 2020 Jun 2;9(11):e013227. DOI: 10.1161/jaha.119.013227

3. Xu Y, Zhao L, Zhang L, Han Y, Wang P, Yu S. Left Atrial Enlargement and the Risk of Stroke: A Meta-Analysis of Prospective Cohort Studies. Front Neurol. 2020;11:26. DOI: 10.3389/fneur.2020.00026

4. Kreimer F, Gotzmann M. Left Atrial Cardiomyopathy - A Challenging Diagnosis. Front Cardiovasc Med. 2022;9:942385. DOI: 10.3389/fcvm.2022.942385

5. Larsen BS, Olsen FJ, Andersen DM, Madsen CV, Møgelvang R, Jensen GB, et al. Left Atrial Volumes and Function, and Long-Term Incidence of Ischemic Stroke in the General Population. J Am Heart Assoc. 2022 Sep 20;11(18):e027031. DOI: 10.1161/jaha.122.027031

6. Vad OB, van Vreeswijk N, Yassin AS, Blaauw Y, Paludan-Müller C, Kanters JK, et al. Atrial cardiomyopathy: markers and outcomes. Eur Heart J. 2025 Oct 15. DOI: 10.1093/eurheartj/ehaf793

7. Deseoe J, Hänsel M, Herzog L, Davoudi N, Luft AR, Gebhard C et al. Left atrial to ventricular volume ratio is a specific marker of atrial cardiopathy. Evidence from the UK Biobank and an ischemic stroke patient cohort. medRxiv; 2025. Available under: https://www.medrxiv.org/content/10.64898/2025.12.18.25342359v1

8. Lim DJ, Varadarajan V, Quinaglia T, Pezel T, Wu C, Noda C, Heckbert SR, Bluemke D, Ambale-Venkatesh B, Lima JAC. Change in left atrial function and volume predicts incident heart failure with preserved and reduced ejection fraction: Multi-Ethnic Study of Atherosclerosis. Eur Heart J Cardiovasc Imaging. 2024;25:1577–1587. doi: 10.1093/ehjci/jeae138

9. Nielsen JB, Kühl JT, Pietersen A, Graff C, Lind B, Struijk JJ, et al. P-wave duration and the risk of atrial fibrillation: Results from the Copenhagen ECG Study. Heart Rhythm. 2015 Sep;12(9):1887–95. DOI: 10.1016/j.hrthm.2015.04.026

10. Schnabel RB, Aspelund T, Li G, Sullivan LM, Suchy-Dicey A, Harris TB, et al. Validation of an atrial fibrillation risk algorithm in whites and African Americans. Arch Intern Med. 2010 Nov 22;170(21):1909–17. DOI: 10.1001/archinternmed.2010.434

11. Maheshwari A, Norby FL, Soliman EZ, Koene R, Rooney M, O’Neal WT, et al. Refining Prediction of Atrial Fibrillation Risk in the General Population With Analysis of P-Wave Axis (from the Atherosclerosis Risk in Communities Study). Am J Cardiol. 2017 Dec 1;120(11):1980–4. DOI: 10.1016/j.amjcard.2017.08.015

12. Kreimer F, Aweimer A, Pflaumbaum A, Mügge A, Gotzmann M. Impact of P-wave indices in prediction of atrial fibrillation-Insight from loop recorder analysis. Ann Noninvasive Electrocardiol. 2021 Sep;26(5):e12854. DOI: 10.1111/anec.12854

13. Xing LY, Diederichsen SZ, Højberg S, Krieger DW, Graff C, Olesen MS, et al. Electrocardiographic markers of subclinical atrial fibrillation detected by implantable loop recorder: insights from the LOOP Study. Europace. 2023 May 19;25(5). DOI: 10.1093/europace/euad014

14. Magnani JW, Zhu L, Lopez F, Pencina MJ, Agarwal SK, Soliman EZ, et al. P-wave indices and atrial fibrillation: cross-cohort assessments from the Framingham Heart Study (FHS) and Atherosclerosis Risk in Communities (ARIC) study. Am Heart J. 2015 Jan;169(1):53–61.e1. DOI: 10.1016/j.ahj.2014.10.009

15. Khurshid S, Friedman S, Reeder C, Di Achille P, Diamant N, Singh P, et al. ECG-Based Deep Learning and Clinical Risk Factors to Predict Atrial Fibrillation. Circulation. 2022 Jan 11;145(2):122–33. DOI: 10.1161/circulationaha.121.057480

16. Yuan N, Duffy G, Dhruva SS, Oesterle A, Pellegrini CN, Theurer J, et al. Deep Learning of Electrocardiograms in Sinus Rhythm From US Veterans to Predict Atrial Fibrillation. JAMA Cardiol. 2023 Dec 1;8(12):1131–9. DOI: 10.1001/jamacardio.2023.3701

17. Christopoulos G, Graff-Radford J, Lopez CL, Yao X, Attia ZI, Rabinstein AA, et al. Artificial Intelligence-Electrocardiography to Predict Incident Atrial Fibrillation: A Population-Based Study. Circ Arrhythm Electrophysiol. 2020 Dec;13(12):e009355. DOI: 10.1161/circep.120.009355

18. Raghunath S, Pfeifer JM, Ulloa-Cerna AE, Nemani A, Carbonati T, Jing L, et al. Deep Neural Networks Can Predict New-Onset Atrial Fibrillation From the 12-Lead ECG and Help Identify Those at Risk of Atrial Fibrillation-Related Stroke. Circulation. 2021 Mar 30;143(13):1287–98. DOI: 10.1161/circulationaha.120.047829

19. Sau A, Sieliwonczyk E, Barker J, Zeidaabadi B, Pastika L, Patlatzoglou K, et al. Prediction of incident atrial fibrillation: A comprehensive evaluation of conventional and artificial intelligence-enhanced approaches. Heart Rhythm. 2025 Aug 22. DOI: 10.1016/j.hrthm.2025.08.024

20. Brant LCC, Ribeiro AH, Eromosele OB, Pinto-Filho MM, Barreto SM, Duncan BB, et al. Prediction of Atrial Fibrillation From the ECG in the Community Using Deep Learning: A Multinational Study. Circ Arrhythm Electrophysiol. 2025 Oct;18(10):e013734. DOI: 10.1161/circep.125.013734

21. Jabbour G, Nolin-Lapalme A, Tastet O, Corbin D, Jordà P, Sowa A, et al. Prediction of incident atrial fibrillation using deep learning, clinical models, and polygenic scores. Eur Heart J. 2024 Dec 7;45(46):4920–34. DOI: 10.1093/eurheartj/ehae595

22. McKeen K, Masood S, Toma A, Rubin B, Wang B. ECG-FM: an open electrocardiogram foundation model. JAMIA Open. 2025 Oct;8(5):ooaf122. DOI: 10.1093/jamiaopen/ooaf122

23. UK Biobank: Protocol for a Large-Scale Prospective Epidemiological Resource, 2007. https://www.ukbiobank.ac.uk/wp-content/uploads/2011/11/UK-Biobank-Protocol.pdf (06 February 2025, date last accessed)

24. Petersen SE, Matthews PM, Francis JM, Robson MD, Zemrak F, Boubertakh R, et al. UK Biobank’s cardiovascular magnetic resonance protocol. J Cardiovasc Magn Reson. 2016 Feb 1;18:8. DOI: 10.1186/s12968-016-0227-4

25. UK Biobank 12-lead (at rest) ECG, 2015 https://biobank.ndph.ox.ac.uk/ukb/ukb/docs/12lead_ecg_explan_doc.pdf (22 November 2025, date last accessed)

26. Bai W, Suzuki H, Huang J, Francis C, Wang S, Tarroni G, et al. A population-based phenome-wide association study of cardiac and aortic structure and function. Nat Med. 2020 Oct;26(10):1654–62. DOI: 10.1038/s41591-020-1009-y

27. Lima EM, Ribeiro AH, Paixão GMM, Ribeiro MH, Pinto-Filho MM, Gomes PR, et al. Deep neural network-estimated electrocardiographic age as a mortality predictor. Nat Commun. 2021 Aug 25;12(1):5117. DOI: 10.1038/s41467-021-25351-7

28. Jordan K, Yaghi S, Poppas A, Chang AD, Mac Grory B, Cutting S, et al. Left Atrial Volume Index Is Associated With Cardioembolic Stroke and Atrial Fibrillation Detection After Embolic Stroke of Undetermined Source. Stroke. 2019 Aug;50(8):1997–2001. DOI: 10.1161/strokeaha.119.025384

29. Tsutsui K, Brimer SB, Ben-Moshe N, Sellal JM, Oster J, Mori H, et al. SHDB-AF: a Japanese Holter ECG database of atrial fibrillation. Sci Data. 2025 Mar 19;12(1):454. DOI: 10.1038/s41597-025-04777-4

30. Huang JY, Yang SH, Gu FS. [Evaluation of 3 chest leads CM5, CC5, and CL5 and factors which influence the results]. Zhonghua Xin Xue Guan Bing Za Zhi. 1989 Apr;17(2):103–6, 27-8.

31. Li J, Aguirre AD, Moura V, Jin J, Liu C, Zhong L, et al. An Electrocardiogram Foundation Model Built on over 10 Million Recordings. Nejm ai. 2025 Jul;2(7). DOI: 10.1056/aioa2401033

32. Tian Y, Li Z, Jin Y, Wang M, Wei X, Zhao L, et al. Foundation model of ECG diagnosis: Diagnostics and explanations of any form and rhythm on ECG. Cell Rep Med. 2024 Dec 17;5(12):101875. DOI: 10.1016/j.xcrm.2024.101875

33. Nguyen HD, Pham T-T, Le N, Nguyen V. TolerantECG: A Foundation Model for Imperfect Electrocardiogram. arXiv preprint arXiv:250709887. 2025.

34. Alonso A, Krijthe BP, Aspelund T, Stepas KA, Pencina MJ, Moser CB, et al. Simple risk model predicts incidence of atrial fibrillation in a racially and geographically diverse population: the CHARGE-AF consortium. J Am Heart Assoc. 2013 Mar 18;2(2):e000102. DOI: 10.1161/jaha.112.000102

35. DeLong ER, DeLong DM, Clarke-Pearson DL. Comparing the areas under two or more correlated receiver operating characteristic curves: a nonparametric approach. Biometrics. 1988 Sep;44(3):837–45.

36. Ho JE, Enserro D, Brouwers FP, Kizer JR, Shah SJ, Psaty BM, Bartz TM, Santhanakrishnan R, Lee DS, Chan C, et al. Predicting Heart Failure With Preserved and Reduced Ejection Fraction. Circulation: Heart Failure. 2016;9:e003116. doi: 10.1161/CIRCHEARTFAILURE.115.003116

37. Camm AJ, Lip GY, De Caterina R, Savelieva I, Atar D, Hohnloser SH, Hindricks G, Kirchhof P. 2012 focused update of the ESC Guidelines for the management of atrial fibrillation: an update of the 2010 ESC Guidelines for the management of atrial fibrillation--developed with the special contribution of the European Heart Rhythm Association. Europace. 2012;14:1385–1413. doi: 10.1093/europace/eus305

38. Havránek tp, Bulková V, Fiala M, Škňouřil L, Chovančík J, Šimek J, et al. Poor relationship between left atrial diameter and volume in patients with atrial fibrillation. Cor et Vasa. 2012 2012/11/01/;54(6):e386-e92. DOI: 10.1016/j.crvasa.2012.11.004

39. Simonyan K, Vedaldi A, Zisserman A. Deep inside convolutional networks: Visualising image classification models and saliency maps. arXiv preprint arXiv:13126034. 2013.

40. Elsheikh S, Hill A, Irving G, Lip GYH, Abdul-Rahim AH. Atrial fibrillation and stroke: State-of-the-art and future directions. Curr Probl Cardiol. 2024 Jan;49(1 Pt C):102181. DOI: 10.1016/j.cpcardiol.2023.102181

41. Alshehri AM. Stroke in atrial fibrillation: Review of risk stratification and preventive therapy. J Family Community Med. 2019 May-Aug;26(2):92–7. DOI: 10.4103/jfcm.JFCM_99_18

42. Kashou AH, May AM, Noseworthy PA. Artificial Intelligence-Enabled ECG: a Modern Lens on an Old Technology. Curr Cardiol Rep. 2020 Jun 19;22(8):57. DOI: 10.1007/s11886-020-01317-x

43. Kornej J, Börschel CS, Benjamin EJ, Schnabel RB. Epidemiology of Atrial Fibrillation in the 21st Century: Novel Methods and New Insights. Circ Res. 2020 Jun 19;127(1):4–20. DOI: 10.1161/circresaha.120.316340

44. Kleindorfer DO, Towfighi A, Chaturvedi S, Cockroft KM, Gutierrez J, Lombardi-Hill D, et al. 2021 Guideline for the Prevention of Stroke in Patients With Stroke and Transient Ischemic Attack: A Guideline From the American Heart Association/American Stroke Association. Stroke. 2021 Jul;52(7):e364–e467. DOI: 10.1161/str.0000000000000375

45. Chollet L, Iqbal SUR, Wittmer S, Thalmann G, Madaffari A, Kozhuharov N, et al. Impact of atrial fibrillation phenotype and left atrial volume on outcome after pulmonary vein isolation. Europace. 2024 Mar 30;26(4). DOI: 10.1093/europace/euae071

46. Lenart-Migdalska A, Kaźnica-Wiatr M, Drabik L, Knap K, Smaś-Suska M, Podolec PP, et al. Assessment of Left Atrial Function in Patients with Paroxysmal, Persistent, and Permanent Atrial Fibrillation using Two-Dimensional Strain. J Atr Fibrillation. 2019 Oct-Nov;12(3):2148. DOI: 10.4022/jafib.2148

47. Marincheva G, Iakobishvili Z, Valdman A, Laish-Farkash A. Left Atrial Strain: Clinical Use and Future Applications-A Focused Review Article. Rev Cardiovasc Med. May 2022;23(5):154.

48. Nadarajah R, Wahab A, Reynolds C, Raveendra K, Askham D, Dawson R, Keene J, Shanghavi S, Lip GYH, Hogg D, et al. Future Innovations in Novel Detection for Atrial Fibrillation (FIND-AF): pilot study of an electronic health record machine learning algorithm-guided intervention to identify undiagnosed atrial fibrillation. Open Heart. 2023;10. doi: 10.1136/openhrt-2023-002447

