## Supplemental material for "Deep learning enables diagnosis of atrial cardiomyopathy from routine 12-lead electrocardiogram"

**Supplementary Material for the Paper: Deep learning enables diagnosis of atrial cardiomyopathy from routine 12-lead electrocardiogram**

### 1. Detailed Methods

#### 1.1 Datasets

##### 1.1.1 UK Biobank

In the UK Biobank, CMR imaging was done using 1.5 T scanners (MAGNETOM Aera, Syngo Platform VD13A, Siemens Healthcare, Erlangen, Germany) according to a standardized protocol [24]. We analyzed data from the first imaging visit. From CMR imaging left atrium maximum volume, left atrium minimum volume and left ventricle end diastolic volume (LVEDV) were previously extracted using deep learning [26] and made available to researchers. LA max and LA min were divided by body surface area to get indexed LA max and indexed LA min. LAEF was calculated by dividing the difference of LA max and LA min by LA max. LALV was calculated by dividing LA max by LVEDV.

The clinical covariates were ascertained as follows. Age was taken as the age at the visit at which the ECG was recorded. For sex, the sex recorded at the initial visit was taken. Ethnicity was taken from the self-report in the touchscreen questionnaire from the initial visit and categorized into White and non-White. Height and weight were measured during the visit at which the ECG was recorded. Blood pressure was measured twice a few moments apart. We averaged the values for our analysis. Smoking status was taken from the touchscreen questionnaire conducted during the visit. Diabetes, heart failure and atrial fibrillation were identified through linked health records using ICD-10 Codes shown in Table S13. The use of antihypertensive drugs was ascertained from the touchscreen questionnaire conducted during the imaging visit. Previous myocardial infarction was ascertained from the algorithmically defined “Date of myocardial infarction” data field.

For calculation of the heart failure risk model, we extracted presence of left bundle branch block and left ventricular hypertrophy from the automated ECG diagnoses of the UKBB based on the ECGs from the second visit also used for evaluation of the deep learning model. Body mass index was also recorded during this visit.

1.1.2. USZ stroke patient cohort

Stroke patients were classified into two groups, an AF group and a non-AF group, in line with previous studies [28]. For a patient to be classified in the AF group, AF had to be either known from medical history or detected during work-up and no other plausible cause of the stroke could be detected during diagnostic work up. To be classified as non-AF, a patient had to have undergone a minimum of 48h of rhythm monitoring without the detection of AF and an alternative, plausible source of stroke had to be identified during diagnostic work-up. Patients fulfilling neither criterion were excluded.

In the USZ stroke cohort the clinical parameters of the CHARGE-AF Score were extracted from the Swiss stroke registry

1.2 Model finetuning

LA max, LA min LAEF and LALV were standardized to give them all similar weight. At test time, predictions were transformed back using mean and standard deviation from the training set. For model finetuning, we first extracted the local representations from the foundation model and performed average pooling, giving us 768 features for each ECG. Based on these features we then build four separate regression heads, one for each predicted imaging feature (LA max, LA min, LAEF and LALV). Each regression head contained two fully connected hidden layers of size 256 and 128 with ReLU activation and one output node.

The ECG-FM model was finetuned using a batch size of 32 with a two-stage training approach. First, only the regression head was trained for 10 epochs using a learning rate of $10^{-3}$, while freezing the model backbone. The regression heads had a dropout rate of 0.3. Then the model backbone was unfrozen and the entire model finetuned with a learning rate of $10^{-5}$ for the backbone and a learning rate of $10^{-3}$ for the regression head. We used a learning rate scheduler, halving the learning rate with a patience of five epochs. We selected the best model checkpoint based on total mean squared error in the validation set and employed early stopping if the model did not improve after 10 epochs. Finetuning was done on a DNA Nexus instance (Type: mem2_ssd2_gpu1_x64, GPU: NVIDIA A10G 24GB) with CUDA version 11.8.0.

The DL-AF model was finetuned with the same parameters, except for the fact that binary cross entropy loss was employed as it is a classification instead of a regression task. It has only one regression head with the same architecture as the regression heads of the previously described model.

1.3 Ascertainment and evaluation of P wave indices (PWI) in the different cohorts.
In the UK Biobank and the primary care cohort we extracted PWI using the full-length ECG. First, we identified P wave onsets, offsets and peaks as well as R onsets using the neurokit function ecg_preprocess() based on lead I. We then removed all cardiac cycles in which any of the described points were missing/not determinable. If less than two complete cycles were identified, the ECG was excluded. Then the PWI were calculated for each cardiac cycle. P offset – P onset was calculated to determine P duration and converted to milliseconds. R onset – P onset was calculated to determine PR Interval. The voltage at P onset in lead II was subtracted from the voltage at P peak in lead II to receive the P amplitude in lead II. Finally, P amplitudes in lead I and lead aVF were calculated in the same manner. The mean of all the described indices was taken for each ECG. To obtain the P axis, the arctangens of the quotient of average P wave amplitude in lead aVF and average P wave amplitude in lead I was taken.

For the USZ stroke patient cohort, PWI were extracted from the ECG metadata. ECGs were recorded on a Schiller AT-102 G2 device and PWI calculated by the clinically validated ECG treatment module installed on the device.

P wave amplitude in lead II, P axis and P duration were then categorized into normal and abnormal according to previous literature using the cutoffs shown in Table S14. PR interval was included in the model as continuous marker. As no widely accepted PWI based model for diagnosis of atrial cardiopathy exists, we investigated the validity of combining these four markers by assessing their association with previous AF in the UK Biobank cohort and with future AF in the Code15% dataset. Even when combining all four parameters in the same model, they were all highly significantly associated with previous and future AF. Results are shown in Table S15 and Table S16. While we cannot exclude the possibility that better P wave indices exist for detecting AtCM, we demonstrate that the ones we selected based on our literature search provide a sensible baseline.

#### 1.4 Exclusion of ECGs showing AF

In all three studied cohorts, ECGs on which AF was present were removed. In the UK Biobank these were identified through the “Automated diagnosis” data field, made available to all UK Biobank researchers. If AF was documented in the automated diagnoses, the ECG was excluded. In the primary care cohort ECGs on which AF was visible were labelled by healthcare workers. In the USZ stroke cohort, we employed a finetuned version of ECG-FM described in [14] which has been previously validated. We only kept ECGs with a below 50% predicted probability of atrial fibrillation and flutter and an above 50% predicted probability of sinus rhythm.

#### 1.5 Segmenting ECGs for determining segment importance

P wave onset, P wave offset, Q peak, S peak, T wave onset and T wave offset were determined using the ecg_preprocess function from neurokit2 based on 5 second ECG segments. Only complete cardiac cycles starting from P wave onset were included in the analysis. The investigated intervals shown in Figure 6 reach from P onset to P offset (P wave), P offset to Q peak (PQ interval), Q peak to S peak (QRS complex), S peak to T onset (ST segment), T onset to T offset (T wave) and T offset to next P onset (TP interval). Importantly these are not identical to the ECG intervals commonly used in clinical practice. This has two reasons. Firstly, we wanted non-overlapping segments and secondly, neurokit2 only provides Q and S peak detection not onset and offset detection, as this is more reliable.

### 2 Supplementary Tables

| | **UKBB: Identifying participants with previous AF episodes** | | | |
|---|---|---|---|---|
| **Model** | **ROC - AUC** | **PR-AUC** | **Brier (%)** | **P - value** |
| Age+ Sex + Pred. indices | **0.722 (0.709-0.735)** | **0.090 (0.079-0.101)** | 2.67 (2.53–2.81) | |
| Age + Sex + PWI | 0.677 (0.664-0.690) | 0.055 (0.050-0.060) | 2.71 (2.57–2.84) | **< 0.001** |
| Age + Sex | 0.666 (0.653-0.679) | 0.049 (0.045-0.054) | 2.71 (2.58-2.85) | **< 0.001** |
| Age + Sex + DL-AF | 0.706 (0.692-0.719) | 0.078 (0.069-0.088) | 2.68 (2.55-2.81) | **< 0.001** |
| | N = 51’202, AF = 1'443 Prevalence: 0.028 | | | |
| | **UKBB: 5-year risk of AF** | | | |
| **Model** | **ROC-AUC** | **PR-AUC** | **Brier (%)** | **P - value** |
| Age+ Sex + Pred. indices | **0.749 (0.728-0.771)** | **0.029 (0.024-0.038)** | **1.29 (1.14-1.45)** | |
| Age + Sex + PWI | 0.718 (0.697-0.740) | 0.018 (0.016-0.021) | 1.30 (1.15-1.46) | **< 0.001** |
| Age + Sex | 0.713 (0.691-0.735) | 0.018 (0.016-0.021) | 1.30 (1.15-1.46) | **< 0.001** |
| Age + Sex + DL-AF | 0.739 (0.717-0.761) | 0.025 (0.020-0.032) | 1.30 (1.14-1.45) | 0.10 |
| | N = 49’759, AF = 300 | Event rate = 0.013 | | |
| | **USZ: Diagnosing patients with AF as cause of stroke** | | | |
| **Model** | **ROC-AUC** | **PR-AUC** | **Brier (%)** | **P - value** |
| Age+ Sex + Pred. indices | **0.725 (0.666-0.785)** | **0.351 (0.255-0.447)** | 1.86 (1.48-2.25) | |
| Age + Sex + PWI | 0.709 (0.650-0.767) | 0.314 (0.232-0.395) | 1.86 (1.46-2.21) | 0.14 |
| Age + Sex | 0.691 (0.629-0.752) | 0.291 (0.215-0.366) | 1.89 (1.49–2.25) | 0.63 |
| Age + Sex + DL-AF | 0.723 (0.664-0.782) | 0.343 (0.246-0.440) | 1.86 (1.46-2.21) | 0.92 |
| | N = 356, AF = 73 | Prevalence: 0.205 | | |

**Table S1**. Model performance on downstream tasks in the UK Biobank and the stroke cohort of models in limited clinical data setting, where only age and sex are available. UKBB: UK Biobank, Pred.: Predicted, DL-AF: deep learning atrial fibrillation, USZ: University Hospital Zurich. ROC-AUC: Area under the receiver operating characteristic curve, PR-AUC: Area under the precision recall curve

| Male Sex | | | |
|---|---|---|---|
| Model | ROC-AUC | PR-AUC | P - value |
| CHARGE-AF + Pred. Indices | 0.742 (0.723 - 0.761) | 0.135 (0.113 - 0.156) | |
| CHARGE-AF + PWI | 0.697 (0.678 - 0.717) | 0.103 (0.085 - 0.121) | < 0.001 |
| CHARGE-AF | 0.691 (0.671 - 0.71) | 0.101 (0.083 - 0.118) | < 0.001 |
| CHARGE-AF + DL | 0.722 (0.703 - 0.741) | 0.123 (0.103 - 0.143) | < 0.001 |
| N = 20’688, AF = 760 | | Prevalence: 0.037 | |

| Female Sex | | | |
|---|---|---|---|
| Model | ROC-AUC | PR-AUC | P - value |
| CHARGE-AF + Pred. Indices | 0.750 (0.726 - 0.774) | 0.082 (0.063 - 0.100) | |
| CHARGE-AF + PWI | 0.724 (0.700 - 0.749) | 0.068 (0.053 - 0.084) | 0.002 |
| CHARGE-AF | 0.725 (0.701 - 0.749) | 0.065 (0.051 - 0.079) | < 0.001 |
| CHARGE-AF + DL | 0.738 (0.714 - 0.762) | 0.080 (0.062 - 0.097) | 0.04 |
| N = 22756, AF = 461 | | Prevalence: 0.020 | |

**Table S2**. Model performance for identifying patients with previous diagnosis of atrial fibrillation in the UK Biobank stratified by sex using CHARGE-AF as base model.

| Male Sex | | | |
|---|---|---|---|
| Model | ROC-AUC | PR-AUC | P - value |
| Age + Sex + Pred. Indices | 0.702 (0.682 - 0.722) | 0.099 (0.085 - 0.114) | |
| Age + Sex + PWI | 0.638 (0.619 - 0.657) | 0.059 (0.052 - 0.066) | < 0.001 |
| Age + Sex | 0.620 (0.6 - 0.639) | 0.053 (0.047 - 0.059) | < 0.001 |
| Age + Sex + DL | 0.674 (0.654 - 0.694) | 0.09 (0.075 - 0.104) | < 0.001 |
| N = 24’200, AF = 889 | | Prevalence: 0.038 | |

| Female Sex | | | |
|---|---|---|---|
| Model | ROC-AUC | PR-AUC | P - value |
| Age + Sex + Pred. Indices | 0.717 (0.694 - 0.741) | 0.065 (0.049 - 0.081) | |
| Age + Sex + PWI | 0.675 (0.651 - 0.699) | 0.043 (0.034 - 0.052) | < 0.001 |
| Age + Sex | 0.673 (0.649 - 0.696) | 0.038 (0.032 - 0.044) | < 0.001 |
| Age + Sex + DL | 0.706 (0.683 - 0.73) | 0.058 (0.045 - 0.071) | 0.12 |
| N = 27’002, AF = 554 | | Prevalence: 0.021 | |

**Table S3**. Model performance for identifying patients with previous diagnosis of atrial fibrillation in the UK Biobank stratified by sex using age + sex as base model.

| Male Sex | | | |
|---|---|---|---|
| Model | ROC-AUC | PR-AUC | P - value |
| CHARGE-AF + Pred. Indices | 0.765 (0.728 - 0.800 | 0.036 (0.027 - 0.054) | |
| CHARGE-AF + PWI | 0.752 (0.716 - 0.785) | 0.027 (0.022-0.036) | 0.28 |
| CHARGE-AF | 0.751 (0.713 - 0.786) | 0.027 (0.022 - 0.037) | 0.21 |
| CHARGE-AF + DL | 0.764 (0.726 - 0.798) | 0.034 (0.026 - 0.051) | 0.91 |
| N = 19’928 | | Event rate = 0.017 | |

| Female Sex | | | |
|---|---|---|---|
| Model | ROC-AUC | PR-AUC | P - value |
| CHARGE-AF + Pred. Indices | 0.781 (0.728 - 0.833) | 0.030 (0.0190.052) | |
| CHARGE-AF + PWI | 0.755 (0.694 - 0.806) | 0.019 (0.0130-0.033) | 0.037 |
| CHARGE-AF | 0.750 (0.692 - 0.805) | 0.018 (0.013 - 0.030) | 0.017 |
| CHARGE-AF + DL | 0.767 (0.711 - 0.814) | 0.019 (0.013 - 0.032) | 0.11 |
| N = 22’295 | | Event rate = 0.0096 | |

**Table S4**. Model performance predicting 5-year risk of atrial fibrillation in the UK Biobank stratified by sex using CHARGE-AF as base model.

| Male Sex | | | |
|---|---|---|---|
| Model | ROC-AUC | PR-AUC | P - value |
| Age + Sex + Pred. Indices | 0.738 (0.703 - 0.777 | 0.031 (0.026 - 0.041) | |
| Age + Sex + PWI | 0.708 (0.665 - 0.740) | 0.020 (0.017– 0.024) | 0.015 |
| Age + Sex | 0.710 (0.671 - 0.745) | 0.020 (0.017 - 0.024) | 0.046 |
| Age + Sex + DL | 0.733 (0.694 - 0.773) | 0.030 (0.024 - 0.040) | 0.70 |
| N = 23’311 | | Event rate = 0.017 | |

| Female Sex | | | |
|---|---|---|---|
| Model | ROC-AUC | PR-AUC | P - value |
| Age + Sex + Pred. Indices | 0.753 (0.698 - 0.799) | 0.027 (0.018–0.058) | |
| Age + Sex + PWI | 0.722 (0.671 - 0.769) | 0.012 (0.010– 0.015) | 0.076 |
| Age + Sex | 0.710 (0.670 - 0.754) | 0.012 (0.009 - 0.015) | 0.011 |
| Age + Sex + DL | 0.740 (0.684 – 0.797) | 0.015 (0.011 - 0.022) | 0. |
| N = 26’448 | | Event rate = 0.010 | |

**Table S5**. Model performance predicting 5-year risk of atrial fibrillation in the UK Biobank stratified by sex using Age + Sex as base model.

| | No Prev. HF | Prev. HF | P-values* |
|---|---|---|---|
| **n** | 42’947 | 323 | |
| **Age (mean (SD))** | 67.66 (7.77) | 72.83 (6.61) | <0.001 |
| **Sex = Male (%)** | 20’381 (47.5) | 226 (70.0) | <0.001 |
| **BMI (mean (SD))** | 26.67 (4.61) | 28.17 (5.32) | <0.001 |
| **Sys. BP [mmHg] (mean (SD))** | 144.22 (19.57) | 142.93 (21.99) | 0.235 |
| **Smoking (%)** | 1337 (3.1) | 7 (2.2) | 0.415 |
| **Antihypertensive medication (%)** | 11’994 (27.9) | 229 (70.9) | <0.001 |
| **Diabetes** | 2699 (6.3) | 46 (14.2) | <0.001 |
| **Previous MI (%)** | 964 (2.2) | 127 (39.3) | <0.001 |
| **LBBB (%)** | 437 (1.0) | 30 (9.3) | <0.001 |
| **LVH (%)** | 919 (2.1) | 12 (3.7) | 0.080 |

**Table S6.** Participant characteristics in the UK Biobank cohort used for evaluation of heart failure associated downstream tasks. Patient with AF were not excluded. Stratification was done according to previous diagnosis of heart failure (HF) being present in health records. Prev: Previous, SD: standard deviation, BMI body mass index, sys. BP: systolic blood pressure, MI: Myorardial infarction, LBBB: left bundle branch block, LVH: Left ventricular hypertrophy.

For age, Body mass index, and systolic blood pressure p-values were calculated using two-tailed unpaired T-test, for the other variables Pearsons chi-squared test was used.

| | **UKBB: Identifying participants with previous diagnosis of HF** | | | |
|---|---|---|---|---|
| **Model** | **ROC - AUC** | **PR-AUC** | **Brier (%)** | **P - value** |
| **HF-Score + Pred. indices** | **0.865 (0.845 - 0.885)** | **0.119 (0.089 - 0.15)** | **0.697 (0.626 - 0.767)** | |
| HF-Score + PWI | 0.85 (0.83 - 0.87) | 0.100 (0.073 - 0.127) | 0.704 (0.633 - 0.776) | **0.002** |
| HF-Score | 0.849 (0.829 - 0.869) | 0.103 (0.074 - 0.132) | 0.702 (0.00634 - 0.00776) | **< 0.001** |
| HF-Score + DL-AF | 0.853 (0.833 - 0.873) | 0.109 (0.08 - 0.138 | 0.700 (0.00633 - 0.763) | **0.025** |
| | N = 43’270, HF = 323 Prevalence: 0.0075 | | | |
| | **UKBB: 5-year risk of HF** | | | |
| **Model** | **ROC-AUC** | **PR-AUC** | **Brier (%)** | **P - value** |
| **HF-Score + Pred. indices** | **0.794 (0.754- 0.833)** | **0.0217 (0.0151 - 0.0420)** | **0.600 (0.409 - 0.719)** | |
| HF-Score + PWI | 0.785 (0.743- 0.827) | 0.00198 (0.0085 - 0.0401 | 0.598 (0.4794 - 0.718) | 0.40 |
| HF-Score | 0.788 (0.747 - 0.830) | 0.0185 (0.0118 - 0.0296) | 0.600 (0.4801 - 0.719) | 0.47 |
| HF-Score + DL-AF | 0.788 (0.746 - 0.830) | 0.0203 (0.0141 - 0.0406) | 0.598 (0.479- 0.717) | 0.42 |
| | N = 42’947, HF = 111 | Event rate = 0.00608 | | |

**Table S7**. Model performance on heart failure related downstream tasks in the UK Biobank. Here participants with previously diagnosed AF were included and participants with incident AF were not censored. P values show results of DeLong’s test for comparing the HF-Score + Pred. Indices models to the other models. UKBB: UK Biobank, HF heart failure, Pred.: Predicted, DL-AF: deep learning atrial fibrillation. ROC-AUC: Area under the receiver operating characteristic curve, PR-AUC: Area under the precision recall curve

| | **UKBB: Identifying participants with previous diagnosis of HF** | | | |
|---|---|---|---|---|
| **Model** | **ROC - AUC** | **PR-AUC** | **Brier (%)** | **P - value** |
| **HF-Score + Pred. indices** | **0.856 (0.829 - 0.884)** | **0.104 (0.069 - 0.139)** | **0.531 (0.464 - 0.595)** | |
| HF-Score + PWI | 0.846 (0.818 - 0.874) | 0.091 (0.057 - 0.124) | 0.535 (0.467 - 0.598) | **0.039** |
| HF-Score | 0.848 (0.82 - 0.876) | 0.095 (0.059 - 0.13) | 0.533 (0.465 - 0.596) | 0.085 |
| HF-Score + DL-AF | 0.849 (0.821 - 0.876) | 0.093 (0.059 - 0.126) | 0.533 (0.467 - 0.597) | 0.082 |
| | N = 42'056, HF = 237 | Prevalence: .0056 | | |
| | **UKBB: 5-year risk of HF** | | | |
| **Model** | **ROC-AUC** | **PR-AUC** | **Brier (%)** | **P - value** |
| **HF-Score + Pred. indices** | **0.779 (0.734- 0.823)** | **0.0149 (0.0087 - 0.0425)** | **0.453 (0.350 - 0.0557)** | |
| HF-Score + PWI | 0.771 (0.727- 0.816) | 0.00145 (0.0085 - 0.0401 | 0.453 (0.349 - 0.556) | 0.40 |
| HF-Score | 0.775 (0.691 - 0.798) | 0.0134 (0.0083 - 0.0341) | 0.453 (0.350 - 0.005567) | 0.47 |
| HF-Score + DL-AF | 0.774 (0.729 - 0.819) | 0.0132 (0.0084 - 0.0307) | 0.453 (0.350- 0.557) | 0.42 |
| | N = 41'819, HF = 87 | Event rate = 0.00458 | | |

**Table S8**. Model performance on heart failure related downstream tasks in the UK Biobank. Here participants with previously diagnosed AF were excluded and participants with incident AF were censored. P values show results of DeLong's test for comparing the HF-Score + Pred. Indices models to the other models. UKBB: UK Biobank, HF heart failure, Pred.: Predicted, DL-AF: deep learning atrial fibrillation. ROC-AUC: Area under the receiver operating characteristic curve, PR-AUC: Area under the precision recall curve

| | aHR | 95%CI | P-value |
|---|---|---|---|
| Pred. LA max | 1.23 | 1.06 - 1.42 | 0.005 |
| Pred. LA min | 1.24 | 1.10 - 1.40 | < 0.001 |
| Pred. LAEF | 0.78 | 0.70 - 0.89 | < 0.001 |
| Pred. LALV | 1.18 | 1.01 - 1.38 | 0.04 |
| N = 42’947 | HF = 152 | | Death = 413 |

**Table S9**. Association of predicted atrial imaging markers with incident heart failure. Separate models were calculated for every predicted imaging marker. Adjusted for parameters shown in Table S6. Participants with previously diagnosed atrial fibrillation were not excluded and participants with incident atrial fibrillation were not censored. The last row shows number of patients reaching the different endpoints. Pred. LA max: predicted left atrial maximum volume indexed to body surface area, Pred. LA min: predicted left atrial minimum volume indexed to body surface area, Pred. LAEF: Predicted left atrial ejection fraction, Pred. LALV: Predicted left atrial to ventricular volume ratio, HR: Hazard ratio, HF heart failure

| | aHR | 95%CI | P-value |
|---|---|---|---|
| Pred. LA max | 1.17 | 1.01 - 1.40 | 0.041 |
| Pred. LA min | 1.22 | 1.06 - 1.41 | 0.006 |
| Pred. LAEF | 0.79 | 0.68 - 0.91 | 0.003 |
| Pred. LALV | 1.12 | 0.93 - 1.34 | 0.23 |
| N = 41’819 | HF = 117 | AF = 268 | Death = 372 |

**Table S10**. Association of predicted atrial imaging markers with incident heart failure. Separate models were calculated for every predicted imaging marker. Adjusted for parameters shown in Table S6. Participants with previously diagnosed atrial fibrillation were excluded and participants with incident atrial fibrillation were censored. The last row shows number of patients reaching the different endpoints. Pred. LA max: predicted left atrial maximum volume indexed to body surface area, Pred. LA min: predicted left atrial minimum volume indexed to body surface area, Pred. LAEF: Predicted left atrial ejection fraction, Pred. LALV: Predicted left atrial to ventricular volume ratio, HR: Hazard ratio, HF heart failure

| Male Sex | | | |
|---|---|---|---|
| Model | ROC-AUC | PR-AUC | P - value |
| Age + Sex + Pred. Indices | 0.828 (0.812 - 0.844) | 0.176 (0.152 - 0.200) | |
| Age + Sex + PWI | 0.755 (0.739 - 0.772) | 0.066 (0.057 - 0.075) | < 0.001 |
| Age + Sex | 0.741 (0.725 - 0.758) | 0.056 (0.049 - 0.062) | < 0.001 |
| Age + Sex + DL | 0.802 (0.786 - 0.819) | 0.194 (0.161 - 0.228) | < 0.001 |
| N = 25’361, AF = 650 | | Prevalence: 0.026 | |

| Female Sex | | | |
|---|---|---|---|
| Model | ROC-AUC | PR-AUC | P - value |
| Age + Sex + Pred. Indices | 0.860 (0.845 - 0.875) | 0.195 (0.163 - 0.227) | |
| Age + Sex + PWI | 0.798 (0.782 - 0.814) | 0.058 (0.053 - 0.064) | < 0.001 |
| Age + Sex | 0.741 (0.725 - 0.758) | 0.051 (0.046 - 0.055) | < 0.001 |
| Age + Sex + DL | 0.843 (0.828 - 0.859) | 0.175 (0.142 - 0.208) | < 0.001 |
| N = 39’430, AF = 605 | | Prevalence: 0.015 | |

**Table S11**. Model performance predicting incident atrial fibrillation in the Brazilian primary care cohort stratified by sex using age + sex as base model.

| Age > 65 years | | | |
|---|---|---|---|
| Marker | ROC-AUC | PR-AUC | P - value |
| Age + Sex + Pred. Indices | 0.765 (0.748 - 0.781) | 0.257 (0.226 - 0.288) | |
| Age + Sex + PWI | 0.622 (0.605 - 0.639) | 0.065 (0.058 - 0.072) | < 0.001 |
| Age + Sex | 0.585 (0.567 - 0.602) | 0.055 (0.05 - 0.06) | < 0.001 |
| Age + Sex + DL | 0.724 (0.706 - 0.742) | 0.207 (0.179 - 0.234) | < 0.001 |
| N = 22’427, AF = 968 | | Prevalence: 0.045 | |

| Age <= 65 years | | | |
|---|---|---|---|
| Marker | ROC-AUC | PR-AUC | P - value |
| Age + Sex + Pred. Indices | 0.800 (0.771 - 0.828) | 0.110 (0.072 - 0.148) | |
| Age + Sex + PWI | 0.727 (0.697 - 0.756) | 0.058 (0.053 - 0.064) | < 0.001 |
| Age + Sex | 0.712 (0.682 - 0.742) | 0.051 (0.046 - 0.055) | < 0.001 |
| Age + Sex + DL | 0.781 (0.75 - 0.811) | 0.109 (0.073 - 0.146) | < 0.001 |
| N = 42’424, AF = 287 | | Prevalence: 0.0067 | |

**Table S12**. Model performance predicting incident atrial fibrillation in the Brazilian primary care cohort stratified by age using age + sex as base model.

| Disease | ICD-10 codes |
|---|---|
| Diabetes | E10 Insulin dependent diabetes mellitus, E11 Non insulin dependent diabetes mellitus, E12 Malnutrition related diabetes mellitus, E13 Other specified diabetes mellitus, E14 unspecified diabetes mellitus |
| Atrial fibrillation/flutter | I48 Atrial fibrillation/flutter |
| Heart failure | I50 heart failure |

**Table S13.** ICD-10 codes used for identification of cardiovascular risk factors

| P wave index | Normal | Abnormal |
|---|---|---|
| P wave amplitude in lead II | ≥ 0.1 mV | < 0.1mv |
| P wave duration | 90 ms – 120 ms | < 90 ms or > 120 ms |
| P wave axis | 0° - 75 ° | < 0 ° or > 75° |

**Table S14.** Classification of P wave indices

| | **Log odds ratio** | **Std. Error** | **P value** |
|---|---|---|---|
| **(Intercept)** | - 5.01 | 0.13 | < 0.001 |
| **Abnormal P amplitude** | 0.31 | 0.10 | **0.002** |
| **Abnormal P Axis** | 0.33 | 0.06 | **< 0.001** |
| **Abnormal P Duration** | 0.19 | 0.06 | **0.002** |
| **PR interval** | 0.008 | 0.0006 | **< 0.001** |
| **N = 49’362, AF = 1’386** | | | |

**Table S15**. Logistic regression model for detecting patients with previous AF episodes in the UK Biobank cohort.

| | **Log odds ratio** | **Std. Error** | **P value** |
|---|---|---|---|
| **(Intercept)** | - 6.02 | 0.10 | < 0.001 |
| **Abnormal P amplitude** | 0.86 | 0.08 | **< 0.001** |
| **Abnormal P Axis** | 0.22 | 0.07 | **0.002** |
| **Abnormal P Duration** | 0.64 | 0.07 | **< 0.001** |
| **PR interval** | 0.010 | 0.0006 | **< 0.001** |
| **N = 64’851, AF = 1’255** | | | |

**Table S16**. Logistic regression model for detecting patients with future AF episodes in the primary care cohort.

### 3. Supplemental Figures

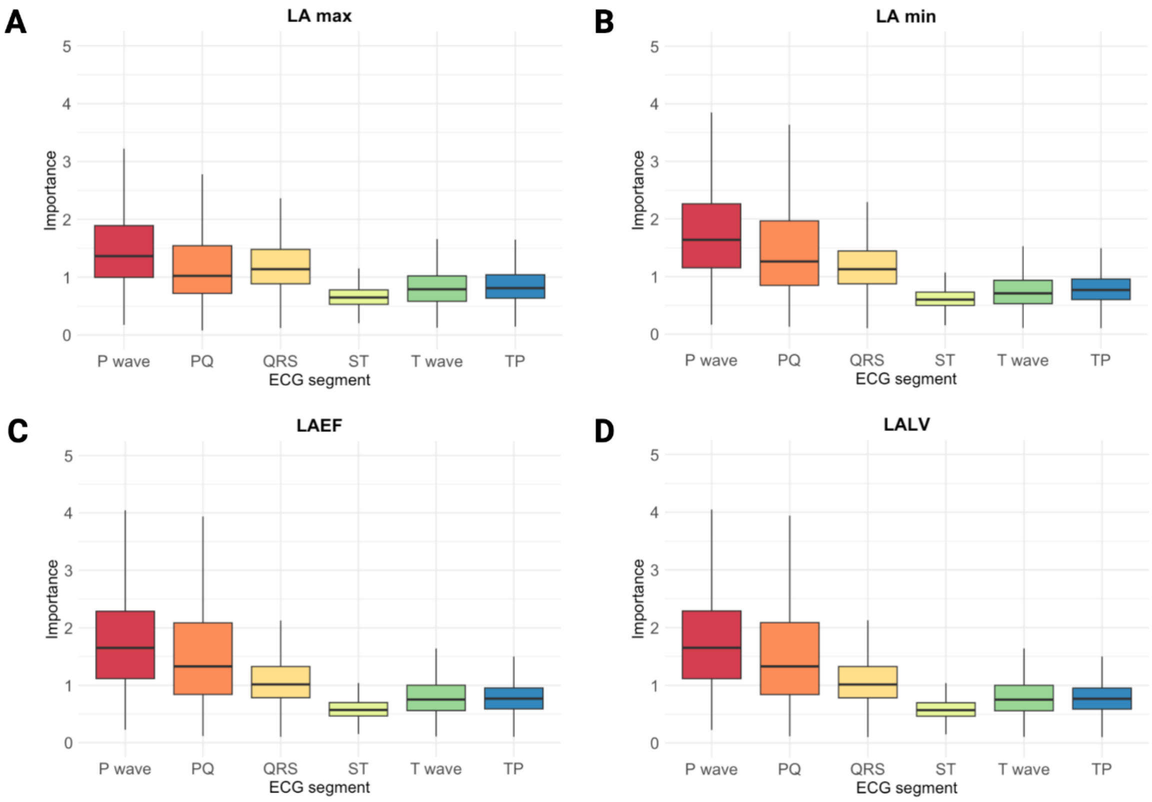


**Figure S1.** Importance of different ECG segments calculated with integrated gradients for prediction of indexed left atrial maximum volume (A), indexed left atrial minimum volume (B), left atrial ejection fraction (C) and left atrial to ventricular volume ratio (D) evaluated on 1'077 5 second non-AF ECG samples from 653 patients from the USZ stroke cohort. Importance was calculated by averaging absolute saliency values over all 12 leads per segment and then dividing by average salience of the entire 5 second ECG sample.
