## Supplemental data Table 1 for "Deep learning enables diagnosis of atrial cardiomyopathy from routine 12-lead electrocardiogram"

**Predicting incident AF**

| Parameters | Pred. Img indices Model | | PWI model | | Clinical Parameter Model | | DL-AF Model | |
|---|---|---|---|---|---|---|---|---|
| | Coefficients | 95%CI | Coefficients | 95%CI | Coefficients | 95%CI | Coefficients | 95%CI |
| Age [years] | 0.0708 | 0.0559 / 0.0856 | 0.0729 | 0.0580 / 0.0879 | 0.0760 | 0.0613 / 0.0907 | 0.0695 | 0.0547 / 0.0844 |
| Sex (male) | 0.274 | 0.028 / 0.521 | 0.444 | 0.227 / 0.661 | 0.472 | 0.259 / 0.686 | 0.383 | 0.217 / 0.593 |
| Pred. LA max [ml / m²] | 0.0541 | 0.0026 / 0.1056 | | | | | | |
| Pred. LA min [ml / m²] | 0.0134 | -0.0665 / 0.0933 | | | | | | |
| Pred. LAEF | -9.14 | -14.89 / -3.39 | | | | | | |
| Pred. LALV | -2.35 | -5.89 / 1.19 | | | | | | |
| Path. P axis | | | 0.107 | -0.114 / 0.327 | | | | |
| Path P.duration | | | 0.159 | -0.080 / 0.398 | | | | |
| Path. P amplitude | | | 0.191 | -0.191 / 0.573 | | | | |
| PR Interval [ms] | | | 0.00110 | -0.00173 / 0.00393 | | | | |
| DL-AF Probability | | | | | | | 18.3 | 14.0 / 22.5 |

**Diagnosing Previous AF**

| Parameters | Pred. Img indices Model | | PWI model | | Clinical Parameter Model | | DL-AF Model | |
|---|---|---|---|---|---|---|---|---|
| | Coefficients | 95%CI | Coefficients | 95%CI | Coefficients | 95%CI | Coefficients | 95%CI |
| Age [years] | 0.0584 | 0.0509 / 0.0658 | 0.0614 | 0.0539 / 0.0689 | 0.0676 | 0.0603 / 0.0749 | 0.0586 | 0.0512 / 0.0661 |
| Sex (male) | 0.444 | 0.319 / 0.570 | 0.434 | 0.323 / 0.544 | 0.534 | 0.426 / 0.642 | 0.401 | 0.291 / 0.512 |
| Pred. LA max [ml / m²] | 0.0252 | -0.0015 / 0.0520 | | | | | | |
| Pred. LA min [ml / m²] | 0.0593 | 0.0203 / 0.0982 | | | | | | |
| Pred. LAEF | -5.25 | -8.06 / -2.44 | | | | | | |
| Pred. LALV | 2.38 | 0.63 / 4.14 | | | | | | |
| Path. P axis | | | 0.188 | 0.076 / 0.300 | | | | |
| Path P.duration | | | 0.102 | -0.019 / 0.222 | | | | |
| Path. P amplitude | | | 0.184 | -0.011 / 0.380 | | | | |
| PR Interval [ms] | | | 0.00563 | 0.00437 / 0.00689 | | | | |
| DL-AF Probability | | | | | | | 21.8 | 19.7 / 24.0 |

**Predicting incident AF**

| Parameters | Pred. Img indices Model | | PWI model | | Clinical Parameter Model | | DL-AF Model | |
|---|---|---|---|---|---|---|---|---|
| | Coefficients | 95%CI | Coefficients | 95%CI | Coefficients | 95%CI | Coefficients | 95%CI |
| Age [years] | 0.0797 | 0.0612 / 0.0982 | 0.0784 | 0.0597 / 0.0972 | 0.0805 | 0.0621 / 0.0989 | 0.0749 | 0.0563 / 0.0934 |
| Race (White) | 0.638 | -0.249 / 1.526 | 0.616 | -0.272 / 1.503 | 0.620 | -0.267 / 1.507 | 0.622 | -0.265 / 1.509 |
| Heart failure | 0.670 | -0.120 / 1.459 | 0.775 | -0.012 / 1.562 | 0.763 | -0.025 / 1.550 | 0.754 | -0.034 / 1.542 |
| Diabetes | 0.104 | -0.301 / 0.509 | -0.00155 | -0.4043 / 0.4012 | 0.0118 | -0.3905 / 0.4140 | 0.0216 | -0.3810 / 0.4242 |
| Smoking (current) | 0.262 | -0.319 / 0.843 | 0.285 | -0.296 / 0.865 | 0.291 | -0.289 / 0.872 | 0.288 | -0.293 / 0.868 |
| Height [cm] | 0.0104 | -0.0050 / 0.0258 | 0.0170 | 0.0027 / 0.0314 | 0.0170 | 0.0028 / 0.0312 | 0.0155 | 0.0012 / 0.0297 |
| Weight [kg] | 0.0195 | 0.0110 / 0.0280 | 0.0198 | 0.0114 / 0.0281 | 0.0203 | 0.0120 / 0.0287 | 0.0177 | 0.0092 / 0.0262 |
| Sys. BP [mmHg] | -0.000641 | -0.00862 / 0.00734 | 0.00398 | -0.00394 / 0.0119 | 0.00393 | -0.00399 / 0.0118 | 0.00341 | -0.00451 / 0.01132 |
| Dias. BP [mmHg] | 0.000822 | -0.0139 / 0.0156 | -0.00822 | -0.02289 / 0.00646 | -0.00790 | -0.02255 / 0.00675 | -0.00642 | -0.02104 / 0.00821 |
| Antihypertensive medication | 0.271 | 0.025 / 0.518 | 0.277 | 0.031 / 0.524 | 0.283 | 0.036 / 0.529 | 0.266 | 0.019 / 0.512 |
| Previous MI | 0.317 | -0.160 / 0.793 | 0.506 | 0.032 / 0.981 | 0.499 | 0.025 / 0.973 | 0.422 | -0.055 / 0.899 |
| Pred. LA max [ml / m²] | 0.0687 | 0.0099 / 0.1275 | | | | | | |
| Pred. LA min [ml / m²] | -0.0141 | -0.1071 / 0.0789 | | | | | | |
| Pred. LAEF | -7.32 | -13.60 / -1.05 | | | | | | |
| Pred. LALV | -1.73 | -5.51 / 2.04 | | | | | | |
| Path. P axis | | | 0.0590 | -0.1800 / 0.2981 | | | | |
| Path P.duration | | | 0.139 | -0.121 / 0.399 | | | | |
| Path. P amplitude | | | 0.170 | -0.254 / 0.594 | | | | |
| PR Interval [ms] | | | -0.000240 | -0.00338 / 0.00290 | | | | |
| DL-AF Probability | | | | | | | 14.3 | 9.3 / 19.2 |

**Diagnosing Previous AF**

| Parameters | Pred. Img indices Model | | PWI model | | Clinical Parameter Model | | DL-AF Model | |
|---|---|---|---|---|---|---|---|---|
| | Coefficients | 95%CI | Coefficients | 95%CI | Coefficients | 95%CI | Coefficients | 95%CI |
| Age [years] | 0.0530 | 0.0436 / 0.0624 | 0.0514 | 0.0419 / 0.0608 | 0.0569 | 0.0476 / 0.0662 | 0.0488 | 0.0394 / 0.0583 |
| Race (White) | 0.529 | 0.117 / 0.941 | 0.497 | 0.091 / 0.904 | 0.520 | 0.114 / 0.926 | 0.486 | 0.077 / 0.895 |
| Heart failure | 1.62 | 1.33 / 1.92 | 1.79 | 1.51 / 2.07 | 1.80 | 1.52 / 2.08 | 1.76 | 1.47 / 2.04 |
| Diabetes | 0.123 | -0.086 / 0.331 | 0.0470 | -0.1572 / 0.2513 | 0.0405 | -0.1631 / 0.2441 | 0.0485 | -0.1575 / 0.2545 |
| Smoking (current) | -0.219 | -0.615 / 0.177 | -0.180 | -0.572 / 0.212 | -0.196 | -0.588 / 0.195 | -0.213 | -0.606 / 0.181 |
| Height [cm] | 0.0331 | 0.0250 / 0.0412 | 0.0293 | 0.0217 / 0.0370 | 0.0327 | 0.0252 / 0.0402 | 0.0310 | 0.0234 / 0.0386 |
| Weight [kg] | 0.00335 | -0.00142 / 0.00812 | 0.00444 | -0.00027 / 0.00916 | 0.00546 | 0.00077 / 0.01014 | 0.00103 | -0.00377 / 0.00583 |
| Sys. BP [mmHg] | -0.00326 | -0.00730 / 0.00079 | 0.00138 | -0.00260 / 0.00536 | 0.00146 | -0.00252 / 0.00543 | 0.000888 | -0.003122 / 0.004898 |
| Dias. BP [mmHg] | -0.0152 | -0.0228 / -0.0076 | -0.0217 | -0.0292 / -0.0142 | -0.0223 | -0.0298 / -0.0148 | -0.0200 | -0.0275 / -0.0124 |
| Antihypertensive medication | 0.508 | 0.379 / 0.637 | 0.521 | 0.393 / 0.648 | 0.526 | 0.399 / 0.654 | 0.495 | 0.366 / 0.624 |
| Previous MI | 0.374 | 0.138 / 0.610 | 0.484 | 0.253 / 0.716 | 0.499 | 0.268 / 0.730 | 0.422 | 0.185 / 0.658 |
| Pred. LA max [ml / m²] | 0.0338 | 0.0037 / 0.0638 | | | | | | |
| Pred. LA min [ml / m²] | 0.0267 | -0.0165 / 0.0700 | | | | | | |
| Pred. LAEF | -3.70 | -6.83 / -0.58 | | | | | | |
| Pred. LALV | 4.30 | 2.42 / 6.18 | | | | | | |
| Path. P axis | | | 0.149 | 0.025 / 0.273 | | | | |
| Path P.duration | | | 0.0748 | -0.0575 / 0.2070 | | | | |
| Path. P amplitude | | | 0.119 | -0.094 / 0.332 | | | | |
| PR Interval [ms] | | | 0.00452 | 0.00310 / 0.00594 | | | | |
| DL-AF Probability | | | | | | | 18.5 | 16.0 / 20.9 |
